# T cell Homeostatic Imbalance in Placentae from Women with HIV in the absence of Vertical Transmission

**DOI:** 10.1101/2021.01.04.21249198

**Authors:** Nadia M. Ikumi, Komala Pillay, Tamara Tilburgs, Thokozile R. Malaba, Sonwabile Dzanibe, Elizabeth Ann L Enninga, Rana Chakraborty, Mohammed Lamorde, Landon Myer, Saye Khoo, Heather B Jaspan, Clive M. Gray, for the DolPHIN-2 Study Group

## Abstract

**Background:** Implementation of universal antiretroviral therapy (ART) has significantly lowered vertical transmission rates but has also increased numbers of HIV-exposed uninfected children (HEU), who remain vulnerable to morbidities. Here, we investigated whether T cell alterations in the placenta contribute to altered immune status in HEU.

**Methods:** We analyzed T cells from term placentae decidua and villous tissue and paired cord blood from pregnant women with HIV (PWH) who initiated ART late in pregnancy (n=21) with pregnant women not living with HIV (PWNH) (n=9).

**Results:** Placentae from PWH showed inverted CD4:CD8 ratios and higher proportions of tissue resident CD8+ T cells in villous tissue relative to control placentae. CD8+ T cells in the fetal capillaries, which were of fetal origin, positively correlated with maternal plasma viraemia prior to ART initiation, implying that imbalanced T cells persisted throughout pregnancy. Additionally, the expanded memory differentiation of CD8+ T cells was confined to the fetal placental compartment and cord blood but was not observed in the maternal decidua.

**Conclusions:** T cell homeostatic imbalance in the blood circulation of PWH is reflected in the placenta. The placenta may be a causal link between HIV-induced maternal immune changes during gestation and altered immunity in newborn infants in the absence of vertical transmission.

**Lay Summary:** The effective prevention of HIV transmission during pregnancy with the rollout of antiretroviral therapy (ART) has resulted in increased numbers of HIV-exposed uninfected children (HEU). These children are vulnerable to infections and health problems and have distorted cellular immune systems at birth. We investigated whether these immune alterations originate in the placenta, as this fetal organ maintains life during pregnancy. After collecting placentae at term from pregnant women living with HIV (PWH), who started ART in the third trimester (n=21) and from pregnant women not living with HIV (PWNH) (n=9), we isolated T cells from dissected placental tissue and matching cord blood. Placentae from PWH showed inverted CD4:CD8 ratios in the placenta and cord blood with higher numbers of CD8+ T cells in the fetal part of the placenta. These CD8+ T cells mirrored events in the blood circulation of the mother and the altered balance of T cell immunity in the PWH was reflected in the placenta. Accordingly, the placenta may be a pivotal link between HIV-induced maternal immune changes and altered immunity in newborn infants in the absence of vertical transmission.

## Background

In adults, HIV causes severe immune dysregulation, characterized by systemic depletion of CD4+ T cells, increased HIV-1 specific CD8+ T cells, inflammation and a progressive failure of the immune system[1–3]. Initiation of antiretroviral therapy (ART) has been shown to augment HIV-specific CD4+ T cell responses, but normalization of the CD4:CD8 T cell ratio does not occur in a large proportion of people with HIV[4]. In pregnant women living with HIV (PWH), there is evidence that women who initiate ART before pregnancy are more likely to have adverse birth outcomes compared to initiation during pregnancy [5].

Placentae from PWH exhibit increased signs of inflammation and injury affecting maternal vasculature and circulation[6,7]. Studies also show that using protease inhibitor-based ART during pregnancy associates with placental injury affecting maternal vascularization and impaired decidualization [8,9]. In addition, although the maternal and fetal circulation within the placenta takes place in distinct compartments, there is evidence that maternal HIV and viral load impacts the fetal immune system. HIV-exposed uninfected children (HEU) have been shown to have lower CD4 T cells and CD4:CD8 T cell ratios at birth [10,11] and this appears to be related to maternal viral loads above 1000 RNA copies/ml [12]. HIV-unexposed uninfected children (HUU) have an almost completely naïve T cell repertoire, but at birth HEU can have increased proportions of differentiated immune cells suggestive of antigen experience *in utero*[10,13]. Indeed, a number of factors including impaired thymic output and functioning may underlie the immune alterations in HEU[14,15]. Here, we sought to investigate how HIV exposure *in utero* may contribute to altered HEU immunity[16,17].

To test the hypothesis that maternal HIV infection is associated with disruption of T cell homeostasis in the placenta and cord blood from HEU newborns, we examined term placentae from PWH from a randomized trial in pregnant mothers initiating dolutegravir versus efavirenz-containing therapy in the third trimester (DolPHIN-2: NCT03249181)[18], as well as pregnant women not living with HIV (PWNH), controls. We show that placentae from PWH have inverted CD4:CD8 ratios with higher CD8+ T cells in villous tissue relative to control placentae contributing to T cell homeostatic imbalance in the placenta at birth.

## Methods

### Cohort

We included 21 placentae with 9 paired cord blood samples from PWH and HEU and 9 placentae from PWNH with 5 cord blood samples from HUU in this study. The PWH group was nested in the DolPHIN-2 study recruited from the Gugulethu Community Health Centre, Cape Town[18]. PWNH were enrolled from Khayelitsha Site B Midwife Obstetric Unit, Cape Town. All placentae were from term deliveries (>37 weeks’ gestation).

### Clinical Data collection

As part of DolPHIN-2, maternal systemic CD4 T cell counts and plasma viral loads (VL) copies were measured at ART initiation at 28 weeks’ gestation (visit 1), 29 weeks (visit 2), 33 weeks (visit 3), 36 weeks (visit 4) and day 14 after delivery (visit 5). The level of HIV-1 RNA detection was 50 copies per ml[18].

### Placenta and cord blood processing

Cells were isolated from each placenta as previously described[19] and illustrated in Supplementary Figure 1. Placentae were collected in RPMI 1640 supplemented with 10% fetal calf serum and penicillin/streptomycin at room temperature and processing was performed within six hours of delivery. Each placenta was dissected to obtain the decidua parietalis, basalis and villous tissue. Enzymatic lymphocyte isolation was performed using Collagenase I and DNAse I. The lymphocyte fraction was obtained following Percoll density centrifugation and incubated with violet amine reactive viability dye (VIVID, Thermofisher). The cells were then fixed using BD FACS ™ lysing solution and cryopreserved in liquid nitrogen until analysis. Cord blood mononuclear cells were isolated on Ficoll, fixed and cryopreserved until analysis.

### Placenta pathology

Whole placentae were fixed in 10% buffered formalin prior to histopathology. Specimens were macroscopically examined and samples from the umbilical cord, placental membranes and placental disk were obtained based on the Amsterdam Placental Workshop Group Consensus Statement[20]. Briefly, four blocks were prepared from each placenta; including a roll of the placental membranes, two cross sections of the umbilical cord; and full-thickness sections of the placental parenchyma and examined in detail as previously described [21].

### Flow cytometry

Placental and cord blood cells were labeled with fluorochrome-conjugated monoclonal antibodies: CD3 (Clone UCHT1), CD4 (Clone SK3), CD8 (Clone SK1), CD45RA (Clone H100), CD28 (Clone CD28.2), CD14 (Clone MHCD1417) and CD45 (Clone MHCD4530). Samples were acquired using an LSR II flow cytometer (BD Biosciences). Total CD4+ and CD8+ T cells were expressed as a proportion of CD3+ T cells (Supplementary Figure 2).

### Immunohistochemistry

Formalin fixed paraffin embedded (FFPE) placenta tissue blocks were cut into 5 µM sections and stained with CD8 (Clone C8/144B), with tonsillar tissue serving as a control. Briefly, the slides were baked overnight at 56°C and rehydrated in xylene followed by varying concentrations of alcohol and then incubated in 3% hydrogen peroxide. Heat-mediated antigen retrieval was performed using an EDTA buffer (pH9). The slides were then incubated with 1% Bovine Serum Albumin and stained with anti-CD8. The images were acquired on Zeiss Axioskop 200 upright Fluorescence microscope with an AxioCam high resolution colour (HRC) camera.

### Fluorescence in situ hybridization (FISH)

As there were five male babies, we selected those placental samples to identify the origin of the infiltrating lymphocytes by looking for the Y chromosome using XY-FISH. Briefly, the slides were baked at 90°C for 15 minutes, deparaffinized in xylene, dehydrated in 100% ethanol and then placed in 10mM Citric Acid (pH 6.0). The slides were then dehydrated in varying concentrations of ethanol (70%, 85% and 100%). We then applied a working solution of DXZ1/DYZ3 (Abbott Laboratories, Des Plaines, IL, USA) to the target areas, co-denatured with a ThermoBrite (Abbott Laboratories) and hybridized overnight at 37°C. The slides were then counter stained with 4’-6’-diamidino-2-phenylindole (DAPI) (Vector Laboratories). Tissue samples were scanned and the qualitative result was determined based on observed signal patterns by CytoVision (Leica Biosystems, Germany).

### Statistics

All flow cytometry data were analyzed using FlowJo version 10 (Treestar). Statistical analyses were performed using Prism version 8 (Graphpad Software), STATA version 12.0 (Stata Corporation) or R[22]. Immunohistochemistry cell counts were performed using Image J Fiji version 2 (WS Rasband, National Institute of Health). Tests of significance were performed using Mann-Whitney *U* and Kruskal-Wallis tests for intergroup comparisons. The associations between cell proportions and maternal viral load or CD4 T cell counts were assessed using simple linear regression. All bivariate analyses including maternal and infant characteristics, and placental pathology stratified by HIV-exposure or by ART regimen were compared using Chi^2^ or Fisher’s exact test and Wilcoxon rank-sum tests.

### Study approval

The study protocol, informed consent and all data collection tools were approved by the University of Cape Town, Human Research Ethics Committee (096/2017). Written and signed informed consent was obtained from all participants, including collection of placentae prior to study inclusion.

## Results

### Participant Characteristics

Maternal and newborn infant characteristics are shown in Table 1. No significant differences in maternal age were noted between PWNH and PWH at enrolment, with the median gestational age (GA) being 30 weeks and 28 weeks between the groups. PWH were more likely to be multigravida (p=0.003). Median GA at delivery was 40 weeks in PWNH and 39 weeks in the PWH (p=0.03) and there was a lower birthweight trend in HEU (p=0.07). We collected placentae from 21 PWH: 16 (76.2%) receiving efavirenz (EFV+TDF+3TC) and 5 (23.8%) receiving dolutegravir (DTG+TDF+3TC), shown in Supplementary Table 1. Median CD4 count at ART initiation was 358 cells/mm^3^ (IQR 278–477), with no difference between ART groups (Supplementary Table 2). The median VL at ART initiation was 4.54 log_10_ RNA copies/ml (IQR 3.85–4.80) in the EFV group versus 3.83 log_10_ RNA copies/ml (3.49– 3.83) in the DTG group, with a combined viraemia of 4.28 log_10_ RNA copies/ml (Supplementary Table 2). Both ART groups were on treatment for a median of 84 days IQR (44-105) and women in the DTG arm achieved viral suppression at a faster rate (cut off ≤ 50 copies/ml or 1.69 log_10_ RNA copies/ml) at 4 weeks versus 2 weeks after delivery (Supplementary Table 2 and [18]). For the purposes of this study, we combined the placentae from two ART groups, as there were only 5 collected from the DTG arm.

**Table 1:** Maternal and infant characteristics.

|  | HIV-Uninfected<br>N=9 | HIV-Infected<br>N=21 | P-Value |
| --- | --- | --- | --- |
| Age, years |  |  | 1.0 |
| ≤24 | 1 (11.1) | 2 (9.5) |  |
| 25-29 | 5 (55.6) | 11 (52.4) |  |
| ≥30 | 3 (33.3) | 8 (38.1) |  |
| Median (IQR) | 29 (25 - 31) | 29 (26 - 30) |  |
| Median gestation at enrolment | 30 (26 - 32) | 28 (28 - 31) | 0.6 |
| Gravidity |  |  | **0.003** |
| 1 | 5 (55.6) | 1 (4.8) |  |
| 2 | 3 (33.3) | 7 (33.3) |  |
| ≥3 | 1 (11.1) | 13 (61.9) |  |
| Median (IQR) | 1 (1 - 2) | 3 (2 - 3) |  |
| **Infant Characteristics** |  |  |  |
| Sex |  |  | 1.0 |
| Female | 6 (66.7) | 12 (57.1) |  |
| Male | 3 (33.3) | 6 (28.6) |  |
| Missing | 0 (0) | 3 (14.3) |  |
| Median gestation at delivery (completed weeks) | 40 (40) | 39 (38 - 39) | **0.003** |
| Birthweight in grams, Median (IQR) | 3420 (3420 - 4000) | 3305 (3010 - 3570) | 0.07 |

### Placental weight is altered by HIV

Table 2 shows placenta characteristics and pathology stratified by HIV status. PWNH had significantly larger placentae (468g, IQR 426-533) compared to the PWH (394g, IQR 343-469; p=0.02), with 38% of placentae in PWH being <10^th^ percentile weight-for-gestation compared to 0% in PWNH. These differences were also reflected the in the fetal-placental (FP) ratios, where all cases with FP ratios >90^th^ percentile were in the PWH cases (p=0.02). Placental histopathology identified 2 cases (9.5%) of chronic deciduitis and 6 cases (28.6%) of MVM, all in the PWH. There were no significant differences in the incidence of meconium exposure and chorioamnionitis between PWH and PWNH, and there was no evidence of villitis of unknown etiology (VUE) in any of the placentae. There were no significant differences in the placental weight, FP ratio and placental pathology between the two ART groups (Supplementary Table 3).

**Table 2:** Placental characteristics and pathology at delivery.

|  | HIV-Uninfected<br>N=9 | HIV-Infected<br>N=21 | P-Value |
| --- | --- | --- | --- |
| Placental basal plate weight (g), median (IQR) | 468 (426 - 533) | 394 (343 - 469) | **0.02** |
| Placental weight (g) |  |  | **0.03** |
| Small (<10<sup>th</sup> percentile) | 0 (0) | 8 (38.1) |  |
| Appropriate (10 - 90<sup>th</sup> percentile) | 9 (100.0) | 12 (57.1) |  |
| Large (>90<sup>th</sup> percentile) | 0 (0) | 0 (0) |  |
| Missing | 0 (0) | 1 (4.8) |  |
| Fetal-Placenta weight ratio |  |  | **0.02** |
| Small (<10<sup>th</sup> percentile) | 1 (11.1) | 0 (0) |  |
| Appropriate (10 - 90<sup>th</sup> percentile) | 8 (88.9) | 11 (52.4) |  |
| Large (>90<sup>th</sup> percentile) | 0 (0) | 7 (33.3) |  |
| Missing | 0 (0) | 3 (14.9) |  |
| Placenta greatest diameter (mm), median (IQR) | 180 (172 - 210) | 180 (170 - 199) | 0.6 |
| Placenta thickness (mm), median (IQR) | 20 (15 -20) | 25 (16.5 - 27.5) | 0.2 |
| Cord insertion |  |  | 0.7 |
| Central | 3 (33.3) | 3 (33.3) |  |
| Off centre | 5 (55.6) | 12 (57.1) |  |
| Marginal | 1 (11.1) | 5 (23.8) |  |
| Missing | 0 (0) | 1 (4.76) |  |
| **Meconium exposure** |  |  | 0.1 |
| + | 3 (33.3) | 2 (9.5) |  |
| - | 6 (66.7) | 19 (90.5) |  |
| **Chorioamnionitis** |  |  |  |
| Maternal Inflammatory Response |  |  | 0.6 |
| + | 2 (22.2) | 2 (9.5) |  |
| - | 7 (77.8) | 19 (90.5) |  |
| Fetal Inflammatory Response |  |  | 0.6 |
| + | 2 (22.2) | 3 (14.3) |  |
| - | 7 (77.8) | 18 (85.7) |  |
| **Chronic Deciduitis** |  |  | 0.5 |
| + | 0 (0) | 2 (9.5) |  |
| - | 9 (100) | 19 (90.5) |  |
| **Maternal Vascular Malperfusion** |  |  | 0.09 |
| + | 0 (0) | 6 (28.6) |  |
| - | 9 (100) | 15 (71.4) |  |
| **Villitis of Unknown Etiology** |  |  | 1.0 |
| + | 0 (0) | 0 (0) |  |
| - | 9 (100) | 21 (100) |  |

### HIV infection during pregnancy alters placental CD4+ and CD8+ T cell proportions

Figure 1A shows significantly lower proportions of CD4+ T cells in decidua parietalis and basalis, but not in the villous tissue, comparing placentae from PWH with PWNH. The proportion of CD8+ T cells was significantly increased (Figure 1A) in all three placental compartments resulting in significantly lower CD4:CD8 T cell ratios in the three placental tissues from PWH (Figure 1B). Notably, the inverted CD4:CD8 ratio in villous tissue was due to the increased CD8+ T cells in the villous tissue. The inverted CD4:CD8 ratio was partially reflected in cord blood from HEU (Figures 1B). No differences were identified in T cell proportions when stratified by the different ART regimens (Supplementary Figure 3). The proportion of CD4+ and CD8+ T cells in the placenta was not associated with placental weight, gravidity or gestational age at delivery (Supplementary Figures 4, 5 and 6).

**Figure 1:**
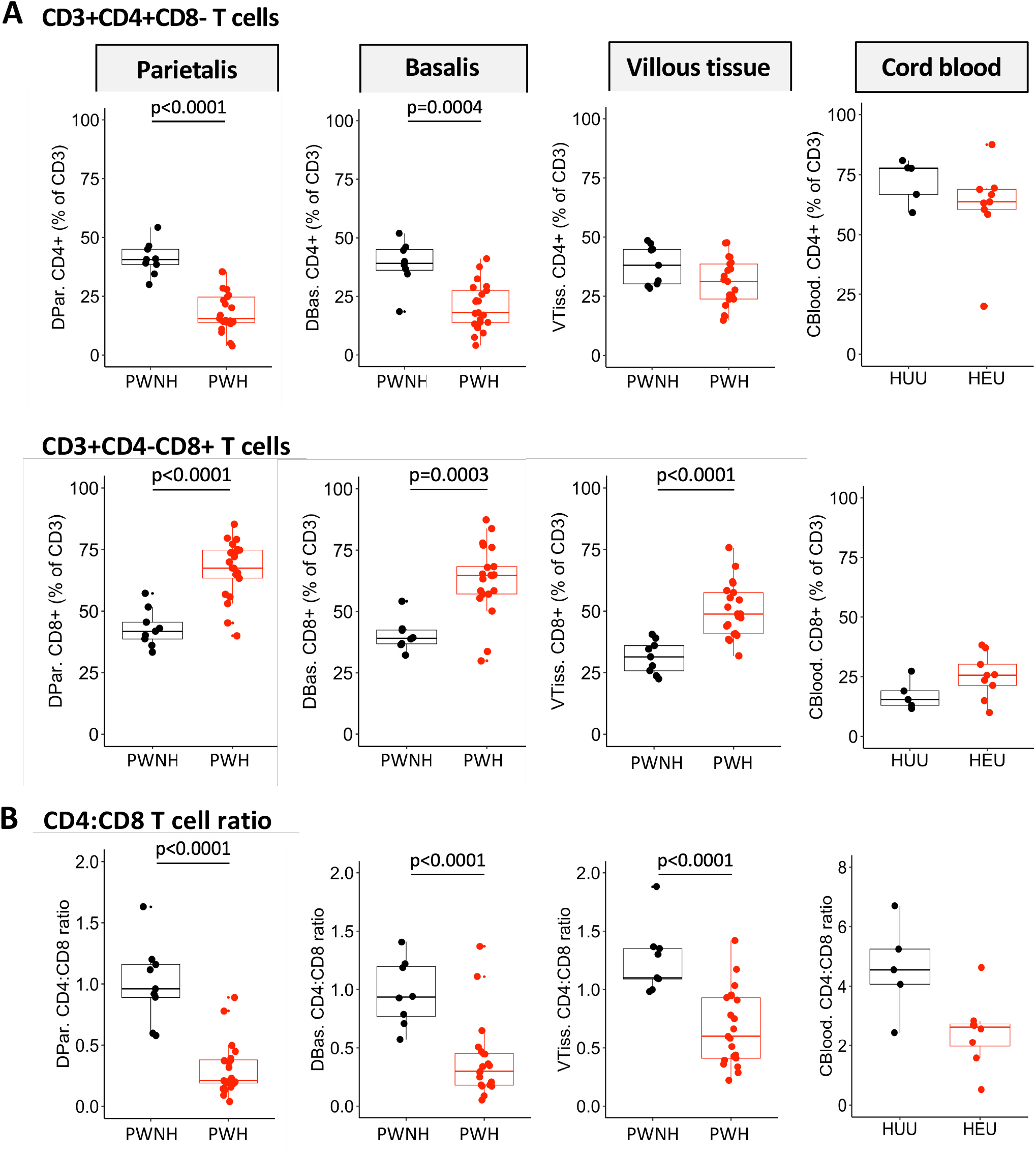
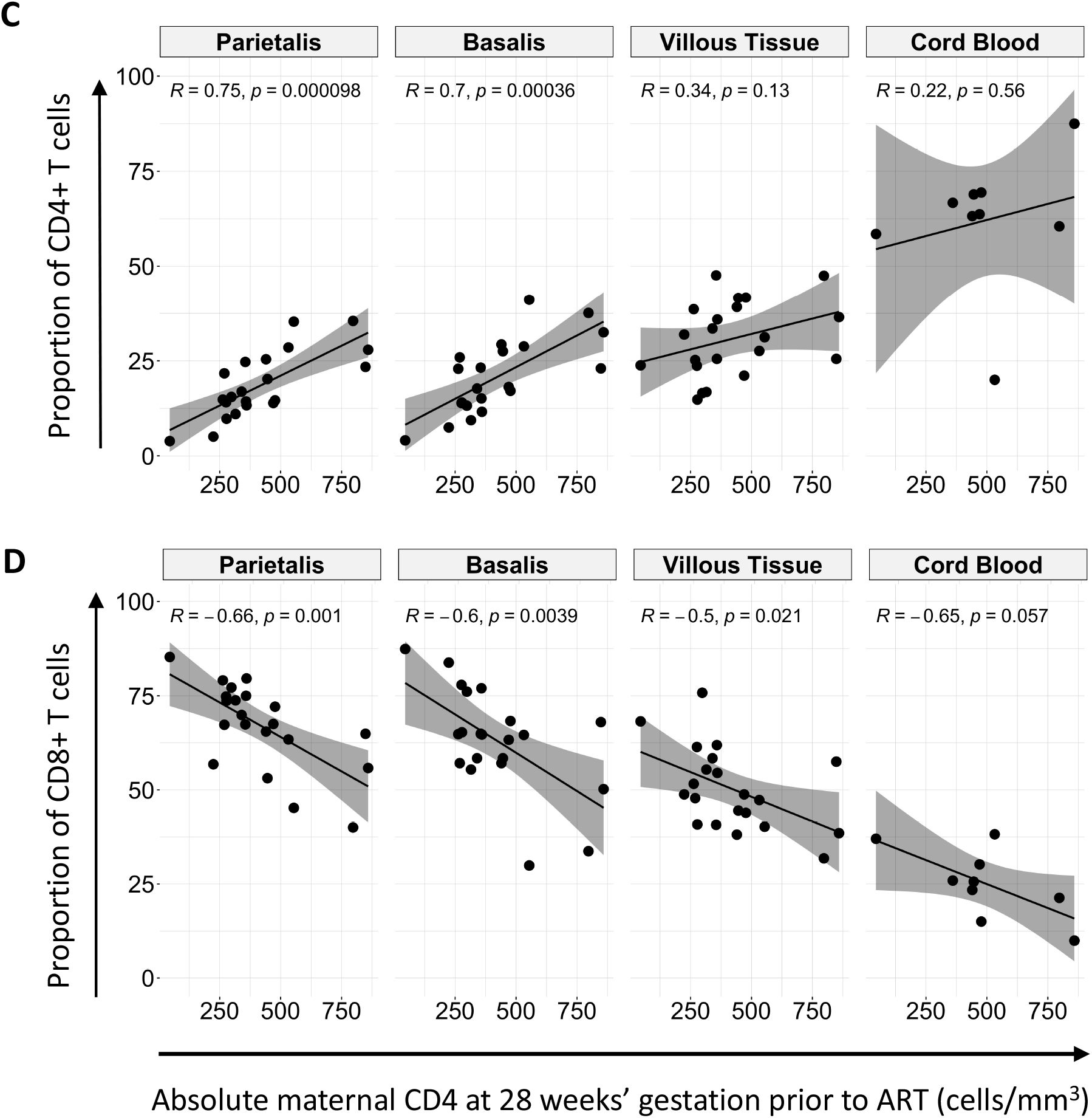
Proportions of T cells in the placenta and cord blood. **(A)** Box plots (showing medians and interquartile ranges) of CD3+CD4+CD8-T cells and CD3+CD4-CD8+ T cell proportions isolated from the decidua parietalis, decidua basalis, villous tissue and cord blood from Pregnant Women not living with HIV (PWNH) and Pregnant Women with HIV (PWH) and HIV unexposed uninfected (HUU) and HIV exposed uninfected (HEU) cord bloods. **(B)** Box plots (showing medians and interquartile ranges) of CD4:CD8 T cell ratios in the decidua parietalis, decidua basalis, villous tissue and cord blood from Pregnant Women not living with HIV (PWNH) and Pregnant Women with HIV (PWH) and HIV unexposed and uninfected (HUU) and HIV exposed uninfected (HEU) cord bloods. Tests of significance were performed using the Mann-Whitney *U* test. **(C)** Correlation plots between the absolute maternal CD4 count at 28 weeks’ gestation prior to ART initiation and the proportion of CD4+ T cells isolated from the decidua parietalis, basalis, villous tissue and cord blood. Statistical analysis was performed using the Spearman rank test and the grey shaded areas represent the 95% confidence intervals. **(D)** Correlation plots between the absolute maternal CD4 count at 28 weeks’ gestation prior to ART initiation and the proportion of CD8+ T cells isolated from the decidua parietalis, basalis, villous tissue and cord blood. Statistical analysis was performed using the Spearman rank test and the grey shaded areas represent the 95% confidence intervals.

Maternal absolute peripheral blood CD4 T cell counts, measured pre-ART at a median of 28 weeks’ gestation, positively correlated with the proportion of CD4+ T cells in the decidua and negatively correlated with the proportion of CD8+ T cells in decidua and villous tissue (Figure 1C and D). A similar trend was observed in cord blood. Thus, the inverted placental tissue CD4:CD8+ T cell ratios appeared to reflect the maternal peripheral immune status but was mirrored to a lesser extent in the cord blood of HEU. This correlation was temporally dissociated, where correlations were made between blood measured at 28 weeks and placentae measured at 38-40 weeks of gestation, suggesting that inverted T cell ratios persisted throughout pregnancy.

### Maternal viral load prior to ART initiation correlates with placental, but not cord blood, CD4+ and CD8+ T cell proportions

Maternal viraemia dropped over time from enrolment to delivery (−12 weeks beforehand), where PWH receiving DTG decreased at a faster rate (Figure 2A)[18]. As expected, the enrolment plasma viraemia, ranging from 1.69 - 6.0 log_10_ RNA copies/ml, significantly inversely correlated with the absolute maternal peripheral blood CD4+ T cell count determined pre-ART, at enrolment (Figure 2B). Pre-ART viremia also showed a significant negative correlation with proportions of CD4+ T cells in the decidua parietalis and basalis (Figure 2C) and a positive correlation with CD8+ T cells in the decidua parietalis, basalis and villous tissue at delivery (Figure 2D). The association between maternal viral load over time and the proportions of decidual CD4+ and CD8+ T cells were maintained at −8 weeks and −4 weeks before delivery for CD4+ T cells (Supplementary Figure 7) and up to −8 weeks before delivery for CD8+ T cells (Supplementary Figure 8). Maternal viremia did not correlate with the proportion of T cells in the cord blood (Figure 2C and 2D), suggesting that maternal viremia (pre- and post-ART initiation) can influence the homeostatic balance of T cells in the placenta, but not in the “newborn” immune compartment.

**Figure 2:**
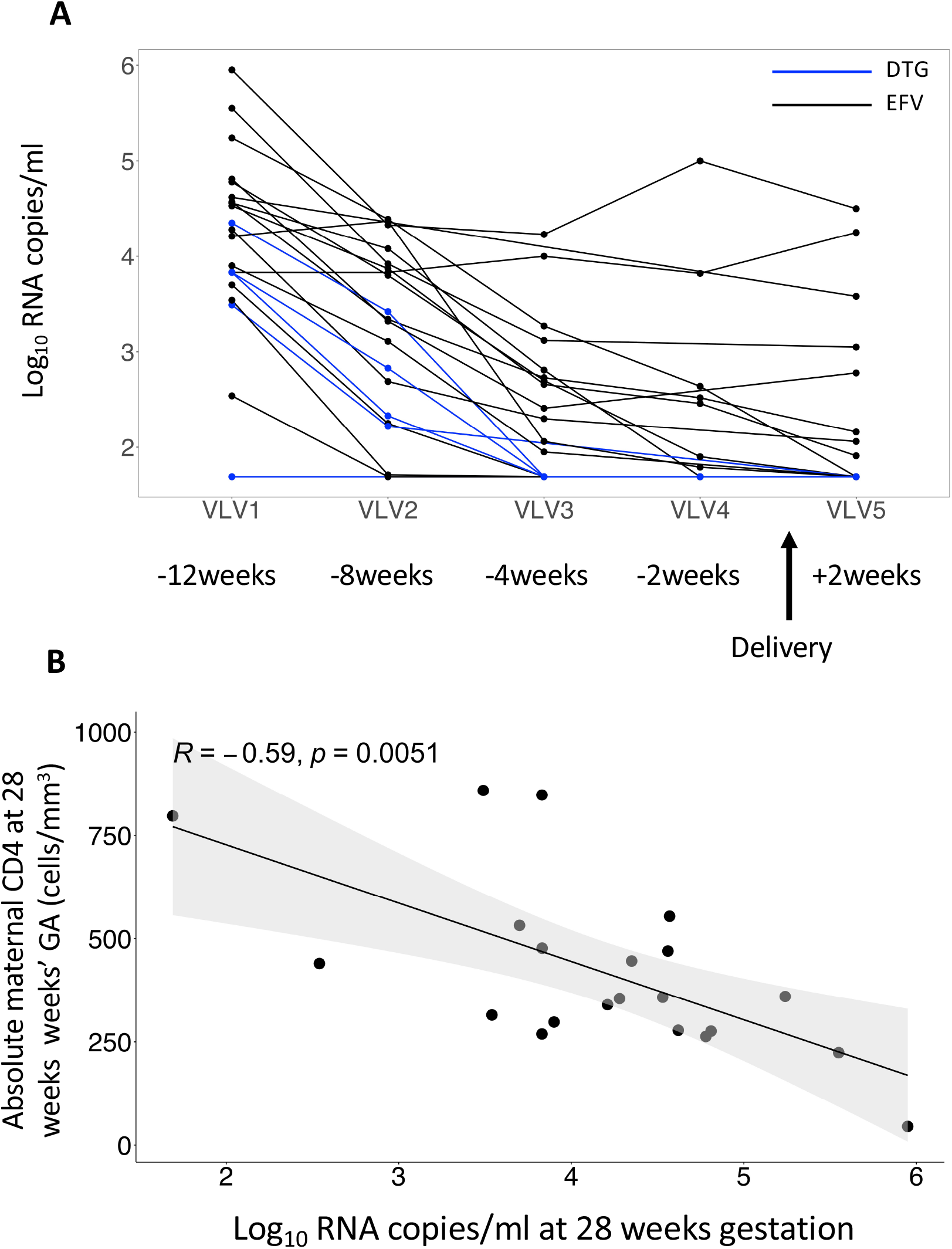
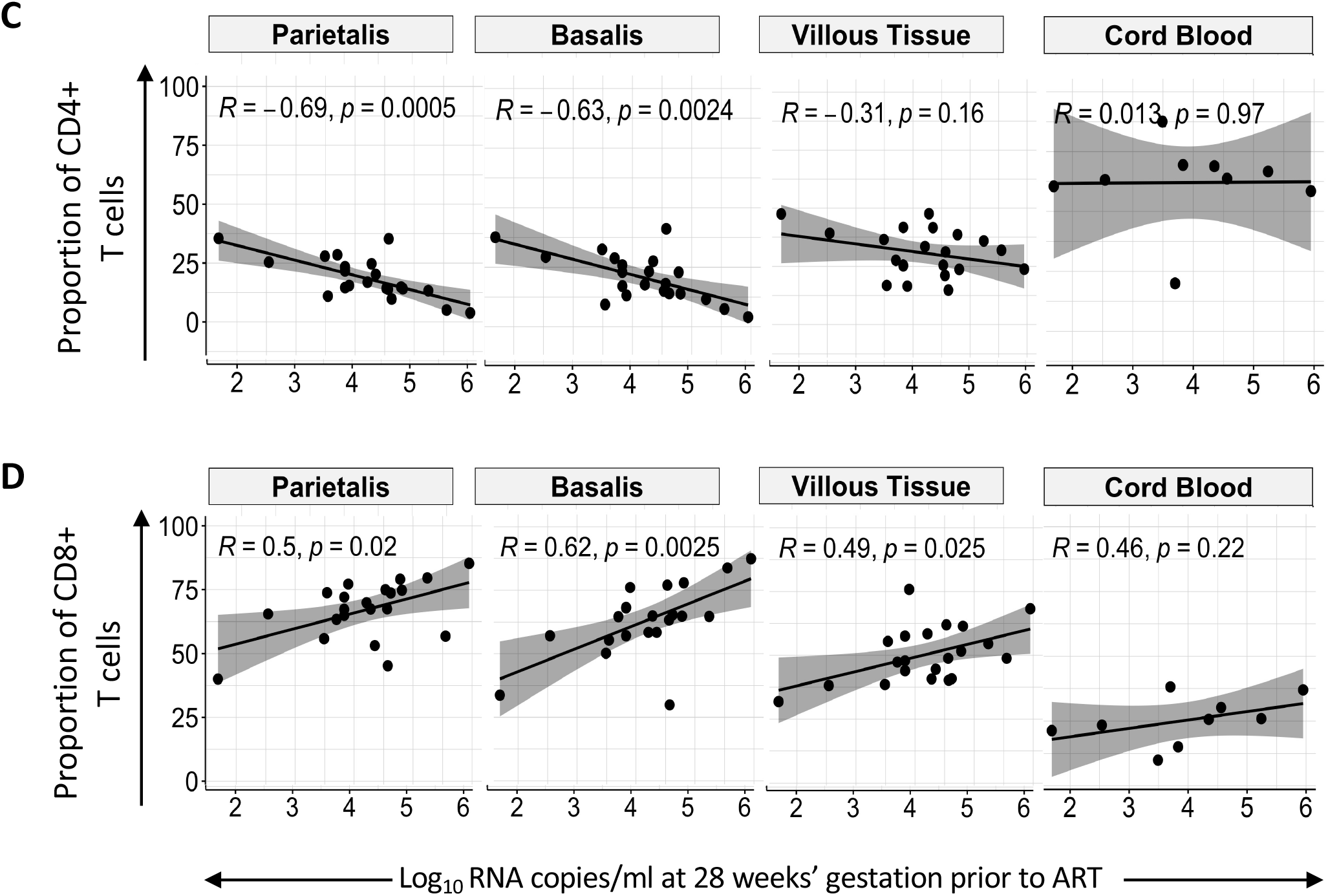
Proportions of CD4+ and CD8+ T cells in the placenta and maternal viral load. **(A)** Line plot depicting participant viral load trajectories over time at ART initiation which was at 28 weeks’ gestation (VLV1, 12 weeks before delivery), 29 weeks’ gestation (VLV2, 8 weeks before delivery), 33 weeks’ gestation (VLV3, 4 weeks before delivery), 36 weeks’ gestation (VLV4, 2 weeks before delivery) and at day 14 after delivery (VLV5, +/-2 weeks after delivery). The women received efavirenz (EFV + TDF + 3TC) (denoted in black) and dolutegravir (DTG + TDF + 3TC) (denoted in blue). **(B)** Correlation plot between maternal systemic absolute CD4 counts at enrolment prior to ART initiation with maternal viral load at enrolment and ART initiation (28 weeks GA). Statistical analysis was performed using the Spearman rank test and the grey shaded area represents the 95% confidence intervals. **(C)** Correlation plots between CD4+ T cell proportions in the placenta and maternal viral load at enrolment and ART initiation (28 weeks GA) in the decidua parietalis, basalis, villous tissue and cord blood. Statistical analysis was performed using the Spearman rank test and the grey shaded area represents the 95% confidence intervals. **(D)** Correlation plots between CD8+ T cell proportions in the placenta and maternal viral load at enrolment and ART initiation (28 weeks GA) in the decidua parietalis, basalis, villous tissue and cord blood. Statistical analysis was performed using the Spearman rank test and the grey shaded area represents the 95% confidence intervals.

### Immunohistochemistry confirms anatomical location and fetal origin of CD8+ T cells in villous tissue

Analysis of placental villi from 13 PWH and 3 PWNH controls showed the presence of CD8+ cells in the fetal capillaries (Figure 3A). The numbers of CD8+ T cells positively correlated with pre-ART maternal viremia (Figure 3B), confirming the flow cytometric analysis (Figures 1A, 2D). Using FISH to detect X and Y chromosomes in cells located within the villi from 5 placentae with male births, we confirmed the presence of male (fetal) cells in fetal capillaries (Figure 3C). Thus, the immunohistochemistry analysis confirms the anatomical location and fetal origin of the increased proportions of CD8+ T cells in villous tissue. The fetal (male) origin of the CD8+ T cells in placental villi is consistent with the absence of villitis of unknown etiology (VUE), a lesion that is characterized by maternal immune infiltrates[23,24].

**Figure 3:**
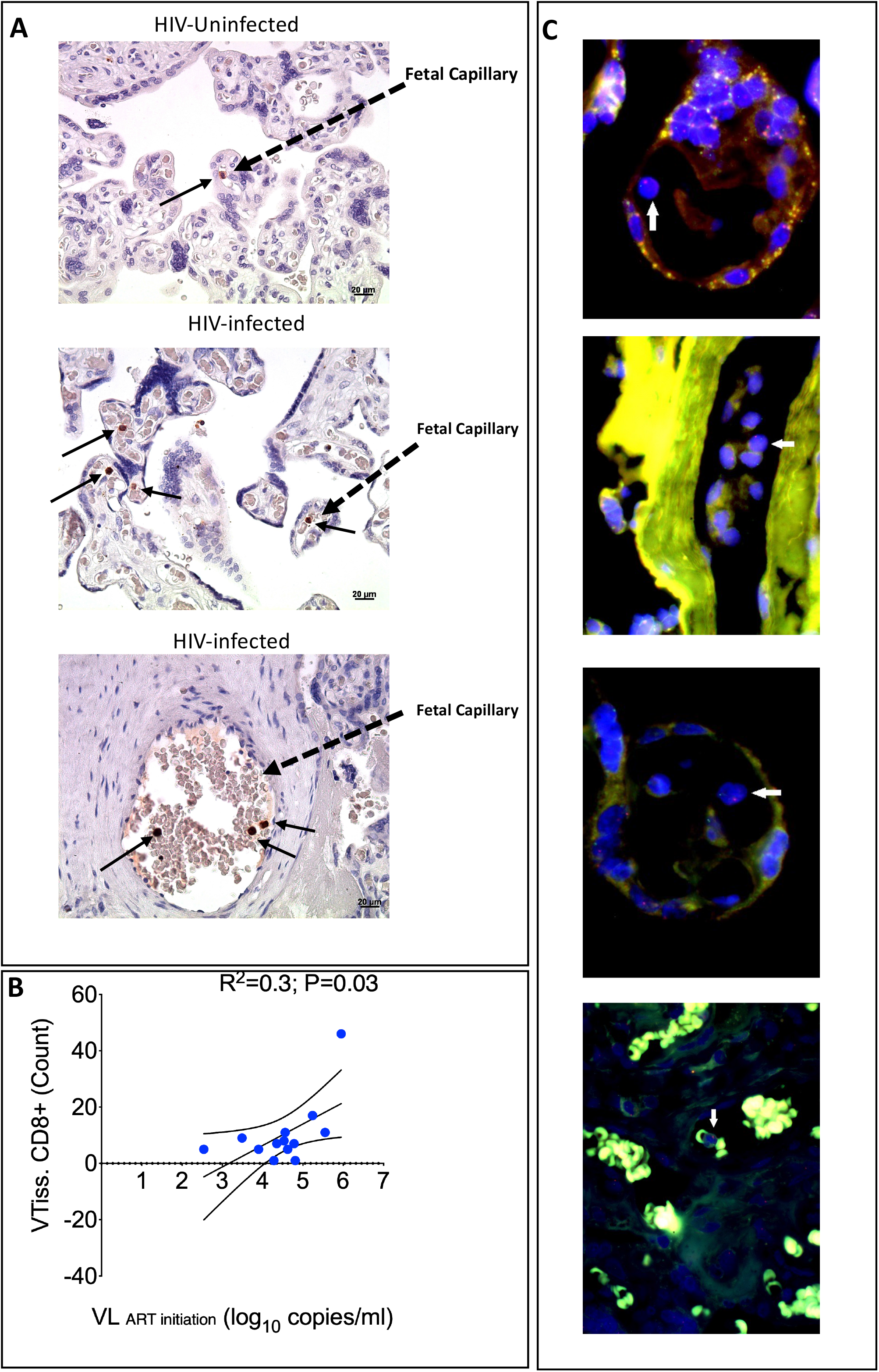
Anatomical location of CD8+ T cells in the villous tissue. **(A)** Representative immunohistochemical stained images of CD8+ T cells in villous tissue sections denoted in brown dots and black arrows in the villi of placentae from HIV-infected and –uninfected mothers (40x magnification). **(B)** Correlation plot between the density of tissue-bound CD8+ T cells in the villi and maternal viral load at ART initiation (pre-ART). Statistical analysis was performed using the Spearman rank test and the black curved lines represent the 95% confidence intervals. **(C)** Representative Fluorescence in situ Hybridization (FISH) images of lymphocytes (white arrows) in the villous tissue from placentas from male infants. The X chromosome is denoted in green and Y chromosome denoted in red (digitally scanned slides).

### HIV exposure increases differentiation of CD8+ T cells in placental villi and fetal cord blood, but not in the maternal placental compartments

CD45RA and CD28 were used to identify the proportions of naïve (CD45RA+CD28+), early differentiated (ED, CD45RA-CD28+), late differentiated (LD, CD45RA-CD28-) and terminally differentiated (TD, CD45RA+CD28-) memory CD4+ and CD8+ T cells from villous tissue and matching cord blood (Figure 4A). We observed significantly lower proportions of naïve CD8+ T cells and significantly higher proportions of LD CD8+ T cells in villous tissue and cord blood of HEU compared to HUU (Figure 4B). The CD4+ T cells in the villous tissue were predominantly of a naïve and ED phenotype while the cord blood cells were predominantly naïve. There were no significant differences in CD4+ T cell differentiation state based on HIV-exposure (Supplementary Figure 9). In addition, no significant differences in the stage of CD4+ and CD8+ T cell differentiation in the decidua was observed between the HIV groups (Supplementary Figure 10). Thus, the increased differentiation state of CD8+ T cells is confined to the fetal placental and cord blood compartments and not observed in the maternal placental compartments. There was no correlation between maternal pre-ART viremia and memory stage of CD8+ T cells in placental villous tissue and cord blood (data not shown). We additionally showed that there was no association between villous tissue CD8+ T cell memory differentiation and reported adverse events in the infant during the first 12 weeks of life (Supplementary Figure 11).

**Figure 4:**
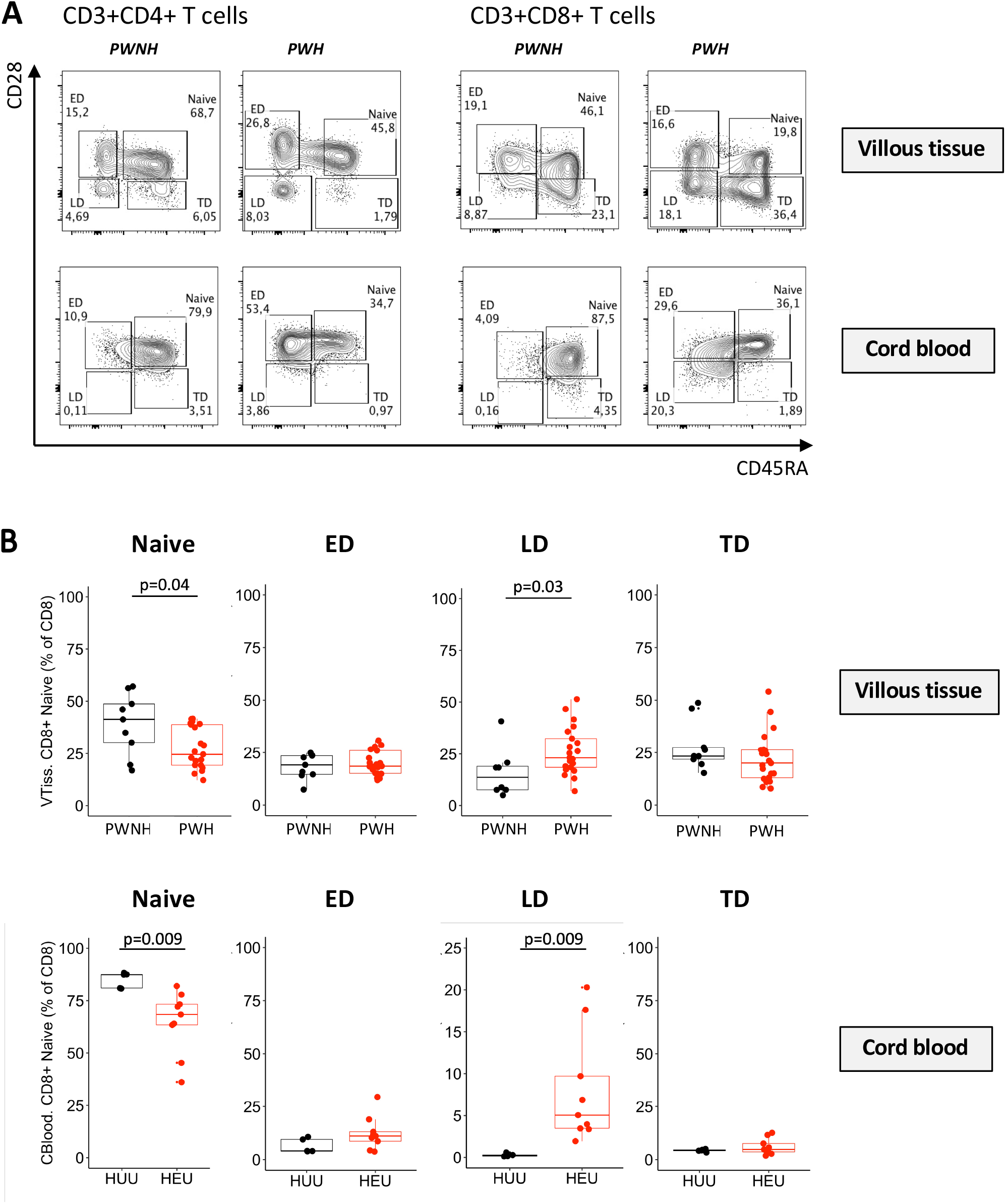
Memory phenotype of CD4 and CD8 T cells in the Villous Tissue. **(A)** Representative flow cytometry contour plots of Naïve, Early differentiated (ED), Late differentiated (LD) and Terminally differentiated (TD) CD4+ T cells and CD8+ T cells in the villous tissue (VT; upper panel) of placentae from PWNH and PWH and cord blood from HIV unexposed and uninfected (HUU) and HIV exposed uninfected (HEU). **(B)** Box plots (showing medians and interquartile ranges) of CD8+ Naïve, Early differentiated (ED), Late differentiated (LD) and Terminally differentiated (TD) T cells in the villous tissue and cord blood from HIV unexposed and uninfected (HUU) and HIV exposed uninfected (HEU). Tests of significance were performed using the Mann-Whitney *U* test.

## Discussion

We present data from a unique cohort of PWH who initiated ART late in pregnancy and show that maternal HIV infection has a clear impact on T cell subsets in the decidua, villous tissue and cord blood. As the decidua and decidual immune cells are of maternal origin, it may not be surprising to find such a footprint[25]. Maternal HIV infection likely affects and kills maternal decidual CD4+ T cells and fewer peripheral blood T cells may traffic to decidual tissue; chemokine gradients have been shown to play a key role in the trafficking of maternal T cells into the decidua during pregnancy[26]. We show that in contrast to the decidua, the inverted CD4:CD8 ratio in the villous tissue was largely due to an increased proportion of CD8+ T cells and not, as observed in the other tissues, due to a decrease in CD4+ T cells. These CD8+ T cells were of an early-late differentiated phenotype, suggestive of previous antigen experience[27,28].

The human placenta has two circulatory compartments: the utero-placental unit for the trafficking of maternal blood and the feto-placental unit for the fetal blood circulation[29,30]. Therefore, cells in the villous tissue are likely to be predominantly from the placental reservoir. A key question is whether the increased CD8+ T cell differentiation in villi and cord blood is due to direct exposure to HIV antigens, presence of other pathogens (e.g. CMV) or increased levels of other non-infectious inflammatory cues in placentae of PWH. Viral particles, structural and core HIV proteins have been shown to cross the placental barrier in the absence of fetal infection leading to altered immune profiles in HEU infants[31,32]. A number of studies also describe HIV-specific T cell responses in HEU infants, possibly primed by exposure to HIV antigens *in utero* [16,33,34].

The lower proportion of naïve cells and increased memory T cells reflected in villous tissue and cord blood mirror previous studies; HEU infants have been shown to have reduced CD4+ T cell numbers and increased CD8+ T cells compared to HUU infants at birth[35,36]. HEU infants have also been shown to have lower naïve cells thought to be due to thymic involution and frequent stimulation and expansion of the antigen-specific T cells in an effort to regenerate the T cell pool[14,15]. Whether the same findings would be recapitulated in PWH with preconception suppressed viral loads is unknown. Nevertheless, this study afforded us a unique opportunity to investigate the impact of starting ART late in pregnancy and how this affected placental immunity. Being able to compare this with women who are consistently on ART throughout pregnancy would reveal whether long-term viral suppression creates a more normalized immune balance in the placenta.

It was not possible to tease out the impact between HIV and ART on the placenta/fetus. There is evidence that some ART can cross the placenta [37,38]. Additional studies show that perinatal drug exposure associates with lower T cell counts in the first two years in HEU infants[39]. Although we cannot discount the effect of ART alone on the placenta, the parent study did report on minimal effects of ART on the newborn child[18]. A limitation of our study was only having maternal CD4 and CD8 T cell counts at week 28 gestation in PWH and no equivalent measures in PWNH. We therefore could not directly compare the impact of HIV/ART exposure on systemic T cells compared to placental T cells at delivery and nor in healthy mothers.

The elevated proportions of CD8+ T cells found embedded in the villi from PWH were shown to be sequestered within the fetal capillaries. These CD8+ T cells within villi were proportional to maternal viremia. The absence of overt VUE corroborates the finding that expanded CD8+ T cell fractions are of fetal and not maternal origin. Previous studies have suggested that the presence of T cells in the villi during normal pregnancy reflect VUE[24]. However, there is emerging evidence, using IHC staining of villi sections from early elective termination placentae, of the presence of CD45+ *αβ* T cells, although these cells were undetectable at term[40]. In a separate study, where there were no reported maternal infections, the villous tissue was shown to contain a mixture of fetal and maternal immune cells[41]. We cannot discount the possibility that there was also a mix of maternal and fetal CD8+ T cells in placental villi in our study.

Here, we demonstrate that within the feto-placental unit, there are differences between the T cell profiles in the villi and cord blood. We postulate that the cells we characterized in the villi may be a combination of cells within the villi, and cells attached to the fetal capillaries. Contaminating circulating cord blood cells in this fraction cannot be completely ruled out but is minimized by extensive washing of the tissues prior to isolation. Moreover, the phenotypic differences between villous and cord blood T cells as presented here suggest that the villous T cells do not include large proportions of circulating T cells. The increased differentiation state of CD8+ T cells in the villi may be due to exposure to antigens within the villous microenvironment as well as the mechanisms of immune activation leading to T cells adhesion and extravasation. Attempts to make a direct connection between the elevated numbers and proportions of CD8+ T cells found in the villous tissue with adverse health events in the first few weeks after birth, revealed no association; most events were resolving rashes.

Of particular note, women were ART naïve during the first and second trimester and it is likely that prolonged HIV exposure may have contributed to altered placental development and the significantly lower placental weight observed. Interestingly, all cases of maternal vascular malperfusion (MVM) were reported in placentae from PWH and possibly reflects placental injury affecting maternal vasculature and perfusion and increasing the risk of an adverse birth outcome[42]. We have previously reported on MVM in placentae from PWH on long-term ART, an incidence of about 27% overall, similar to Kalk *et al* [6,21]. It is likely that HIV and/or ART exposure alters factors involved in vascular development, resulting in placental insufficiency and increased risk of adverse birth outcomes[8,9,43].

In conclusion, we provide evidence that *in utero* exposure to HIV results in an altered immune balance in both the utero-placental and feto-placental compartments. Despite the initiation of ART in the third trimester, resulting in either full or partial maternal viral suppression by the time of delivery, there was a significant imbalance in term placental T cell homeostasis and to a lesser degree in the cord blood. Overall, our results suggest that maternal immunity leaves a footprint in the placenta that may shape the neonatal/infant immune landscape.

## Data Availability

All data that was used in this manuscript is available from Clive Gray

## Author contributions

NMI, ML, SK, LM, HBJ, CMG: Conceptualization and design of the study

NMI, TT: Panel design, sample preparation and development of methods

KP: Histopathology scoring and interpretation

NMI, TRM and SD: Statistical analysis

NMI, TT, KP, TM, LM, EAE, RC, HBJ, CMG: Writing the manuscript

## Acknowledgments

We wish to thank all the study participants in this study; all the members of the DOLPHIN-2 clinical trial and INFANT placenta study. We also wish to thank Nonzwakazi Bangani, Goitseone Thamae, Michelle Barboure, Berenice Alinde and Lizette Fick for their expert assistance in sample processing and Dr Amsha Ramburan for capturing the FISH images.

## Supplementary Figures

**Supplementary Figure 1: Lymphocyte isolation from the human placenta** Stepwise isolation of lymphocytes from the human placenta decidua parietalis (DP), decidua basalis (DB) and villous tissue (VT): (1) dissection of the whole placenta; (2) multiple rounds of maternal blood rinsing to avoid contamination followed by enzymatic digestion of each dissected tissue; (3) lymphocyte separation obtained by Percoll density centrifugation; (4) cryopreservation of fixed cells and immunophenotyping using flow cytometry. Image created with Biorender.com.

**Supplementary Figure 2: Gating strategy to delineate CD3+ T cells** Representative flow cytometry dot plots depicting the gating strategy to delineate CD4+ and CD8+ T cells gated from a live CD45+ CD14-CD3+ population in the placenta.

**Supplementary Figure 3: Proportions of T cells in the placenta and cord blood by ART group (A)** Box plots (showing medians and interquartile ranges) of CD3+CD4+CD8-T cells and CD3+CD4-CD8+ T cell proportions isolated from the decidua parietalis, decidua basalis, villous tissue and cord blood from Pregnant Women not living with HIV (PWNH) and Pregnant Women with HIV (PWH) and HIV unexposed uninfected (HUU) and HIV exposed uninfected (HEU) cord bloods. **(B)** Box plots (showing medians and interquartile ranges) of CD4:CD8 T cell ratios in the decidua parietalis, decidua basalis, villous tissue and cord blood from Pregnant Women not living with HIV (PWNH) and Pregnant Women with HIV (PWH) and HIV unexposed and uninfected (HUU) and HIV exposed uninfected (HEU) cord bloods. Tests of significance were performed using the Mann-Whitney *U* test.

**Supplementary Figure 4: Proportions of T cells in the placenta and cord blood stratified by placental weight** (A) Histogram (showing medians and interquartile ranges) of CD3+CD4+CD8-T cells, (B) CD3+CD4-CD8+ T cells proportions and (C) CD4:CD8 T cell ratios stratified by placental weight in placentas from Pregnant Women not living with HIV (PWNH) and Pregnant Women with HIV (PWH). Tests of significance were performed using the Kruskal-Wallis test.

**Supplementary Figure 5: Proportions of T cells in the placenta and cord blood stratified by gravidity** (A) Histogram (showing medians and interquartile ranges) of CD3+CD4+CD8-T cells, (B) CD3+CD4-CD8+ T cells proportions and (C) CD4:CD8 T cell ratios stratified by gravidity in placentas from Pregnant Women not living with HIV (PWNH) and Pregnant Women with HIV (PWH). Tests of significance were performed using the Kruskal-Wallis test.

**Supplementary Figure 6: Proportions of T cells in the placenta and cord blood stratified by gestational age at delivery** (A) Histogram (showing medians and interquartile ranges) of CD3+CD4+CD8-T cells, (B) CD3+CD4-CD8+ T cells proportions and (C) CD4:CD8 T cell ratios stratified by gestational age at delivery in placentas from Pregnant Women not living with HIV (PWNH) and Pregnant Women with HIV (PWH). Tests of significance were performed using the Kruskal-Wallis test.

**Supplementary Figure 7: Proportions of CD4+ T cells in the placenta, cord blood and maternal viral load** Correlation plots between CD4+ T cell proportions in the placenta and cord blood and maternal viral load at 29 weeks’ gestation (VLV2, 8 weeks before delivery), 33 weeks’ gestation (VLV3, 4 weeks before delivery) and 36 weeks’ gestation (VLV4, 2 weeks before delivery) in the decidua parietalis, basalis, villous tissue and cord blood. Statistical analysis was performed using the Spearman rank test and the grey shaded area represents the 95% confidence intervals.

**Supplementary Figure 8: Proportions of CD8+ T cells in the placenta, cord blood and maternal viral load** Correlation plots between CD8+ T cell proportions in the placenta and cord blood and maternal viral load at 29 weeks’ gestation (VLV2, 8 weeks before delivery), 33 weeks’ gestation (VLV3, 4 weeks before delivery) and 36 weeks’ gestation (VLV4, 2 weeks before delivery) in the decidua parietalis, basalis, villous tissue and cord blood. Statistical analysis was performed using the Spearman rank test and the grey shaded area represents the 95% confidence intervals.

**Supplementary Figure 9: Memory differentiation of CD4+ T cells in the villous tissue and cord blood** Box plots (showing medians and interquartile ranges) of CD4+ Naïve, Early differentiated (ED), Late differentiated (LD) and Terminally differentiated (TD) T cells in the villous tissue (VT; upper panel) and cord blood (CB; lower panel) from HIV-uninfected (MHIV neg) and infected mothers (MHIV pos) and HIV unexposed and uninfected (HUU) and HIV exposed uninfected (HEU) cord bloods. Tests of significance were performed using the Mann-Whitney *U* test.

**Supplementary Figure 10: Memory differentiation of CD4 and CD8 T cells in the decidua parietalis and decidua basalis (A)** Representative flow cytometry contour plots of Naïve, Early differentiated (ED), Late differentiated (LD) and Terminally differentiated (TD) CD4+ T cells and CD8+ T cells in the decidua parietalis and basalis of placentae from PWNH and PWH. **(B)** Box plots (showing medians and interquartile ranges) of CD4+ and **(C)** CD8+ Naïve, Early differentiated (ED), Late differentiated (LD) and Terminally differentiated (TD) T cells in the decidua parietalis and basalis of placentae from PWNH and PWH.

**Supplementary Figure 11: Proportion of CD8+ T cells in the Villous Tissue and Adverse Events in HEU in the first three months of life**. Scatter plot showing the proportion of CD8+ T cells delineated by naïve, early differentiated memory, late differentiated and terminally differentiated memory in HEU infants comparing adverse events with no events reported.

