## Supplemental Figures and Table for "T cell Homeostatic Imbalance in Placentae from Women with HIV in the absence of Vertical Transmission"

### Slide 1
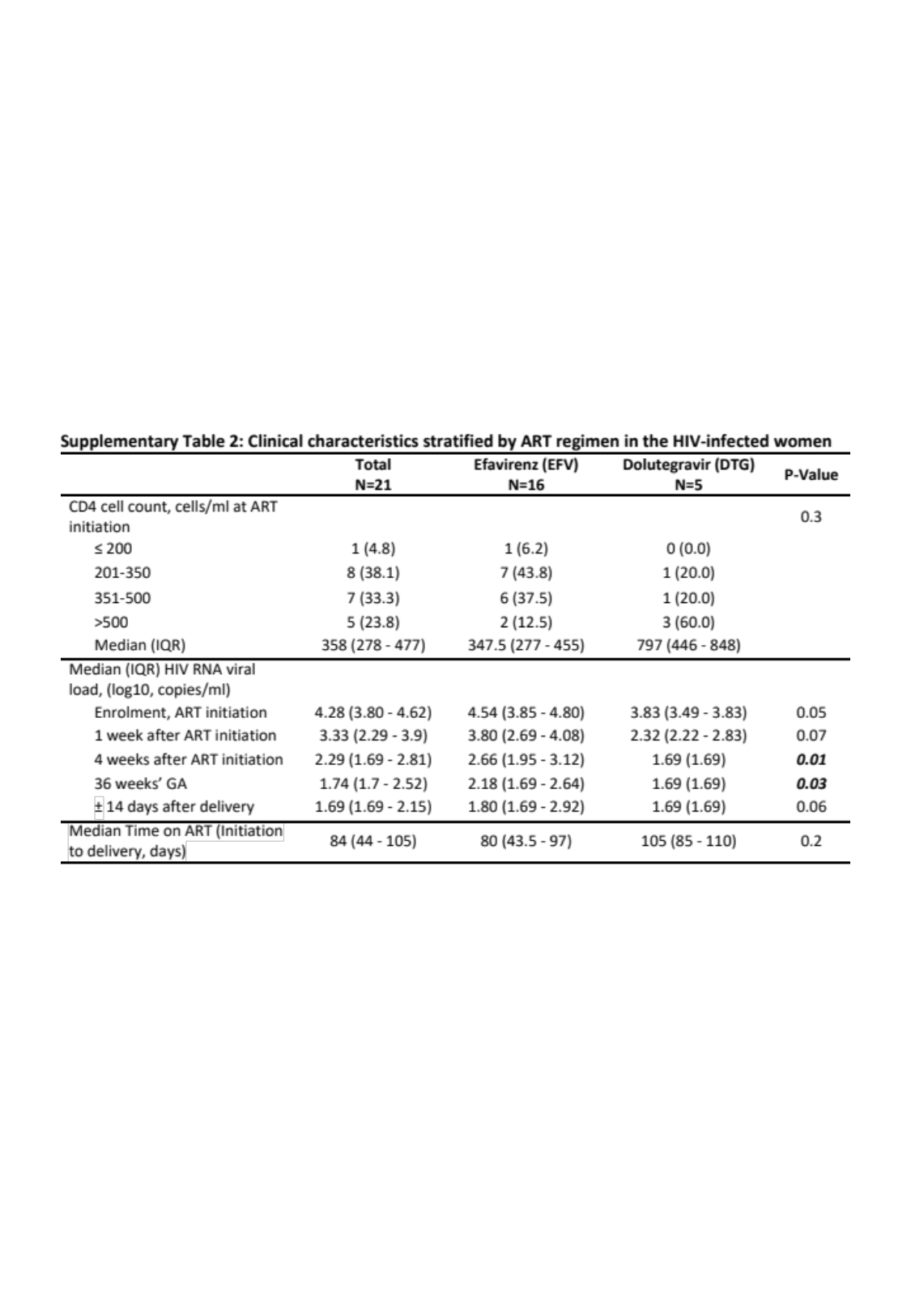

### Slide 2
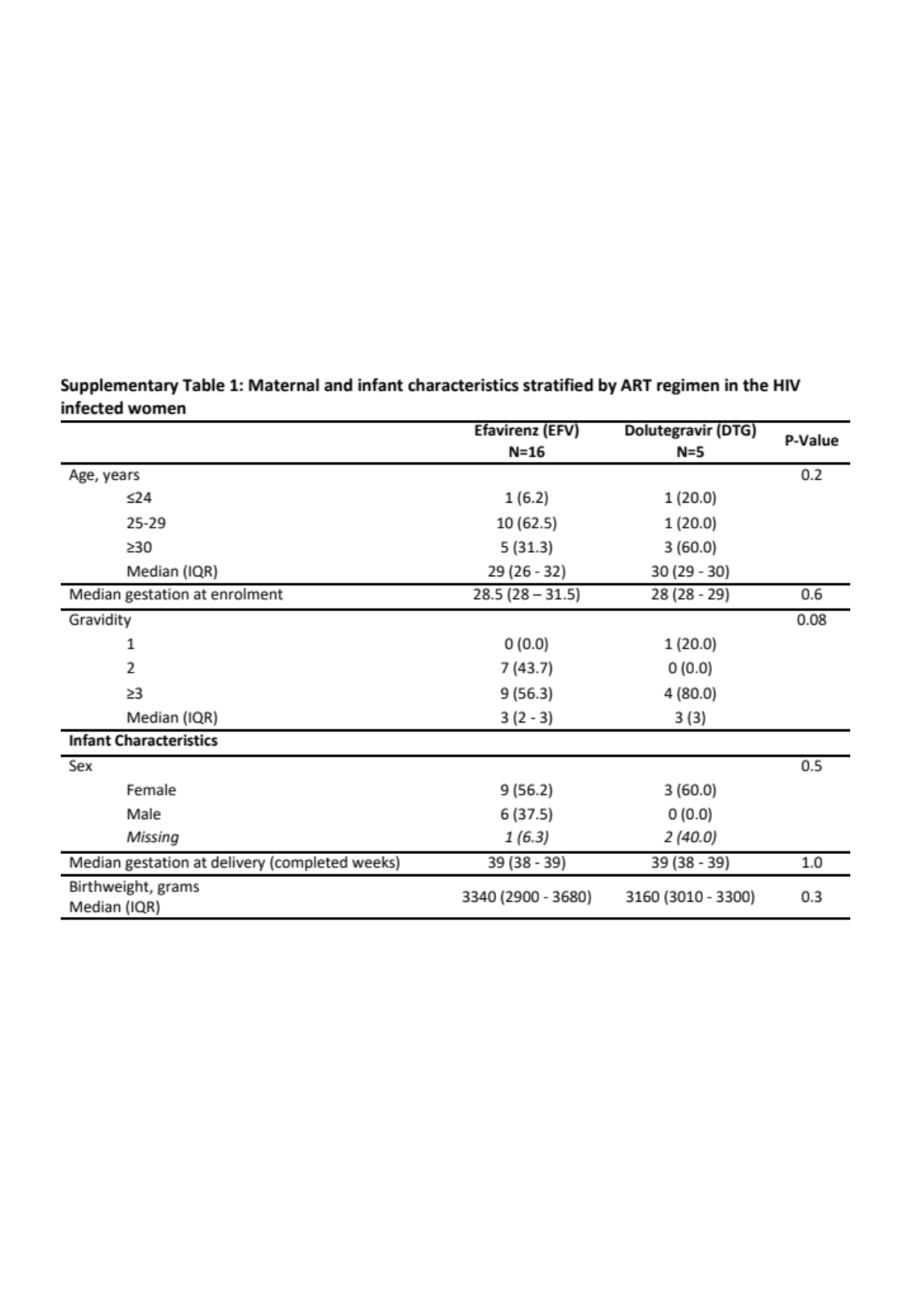

### Slide 3
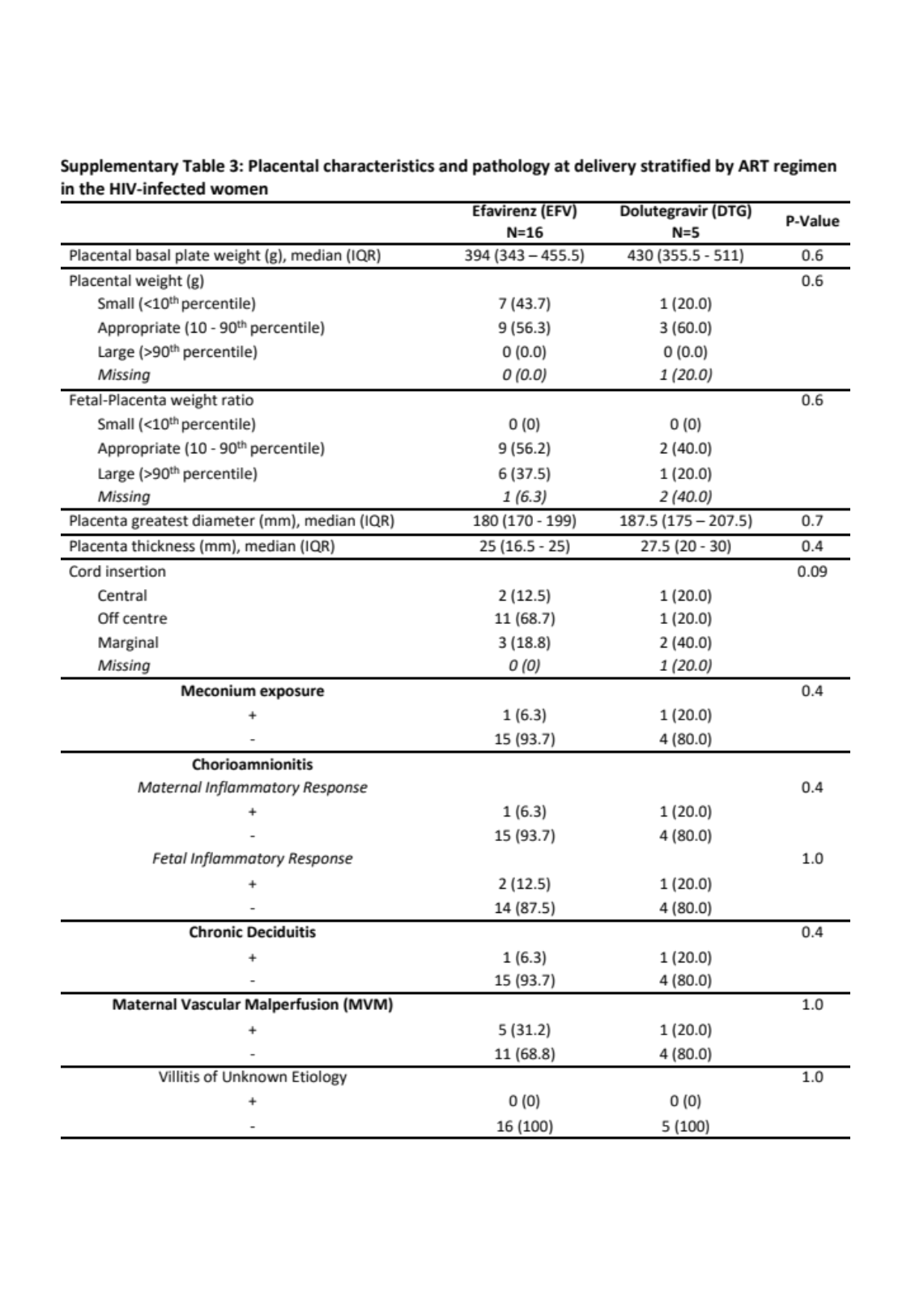

### Slide 4
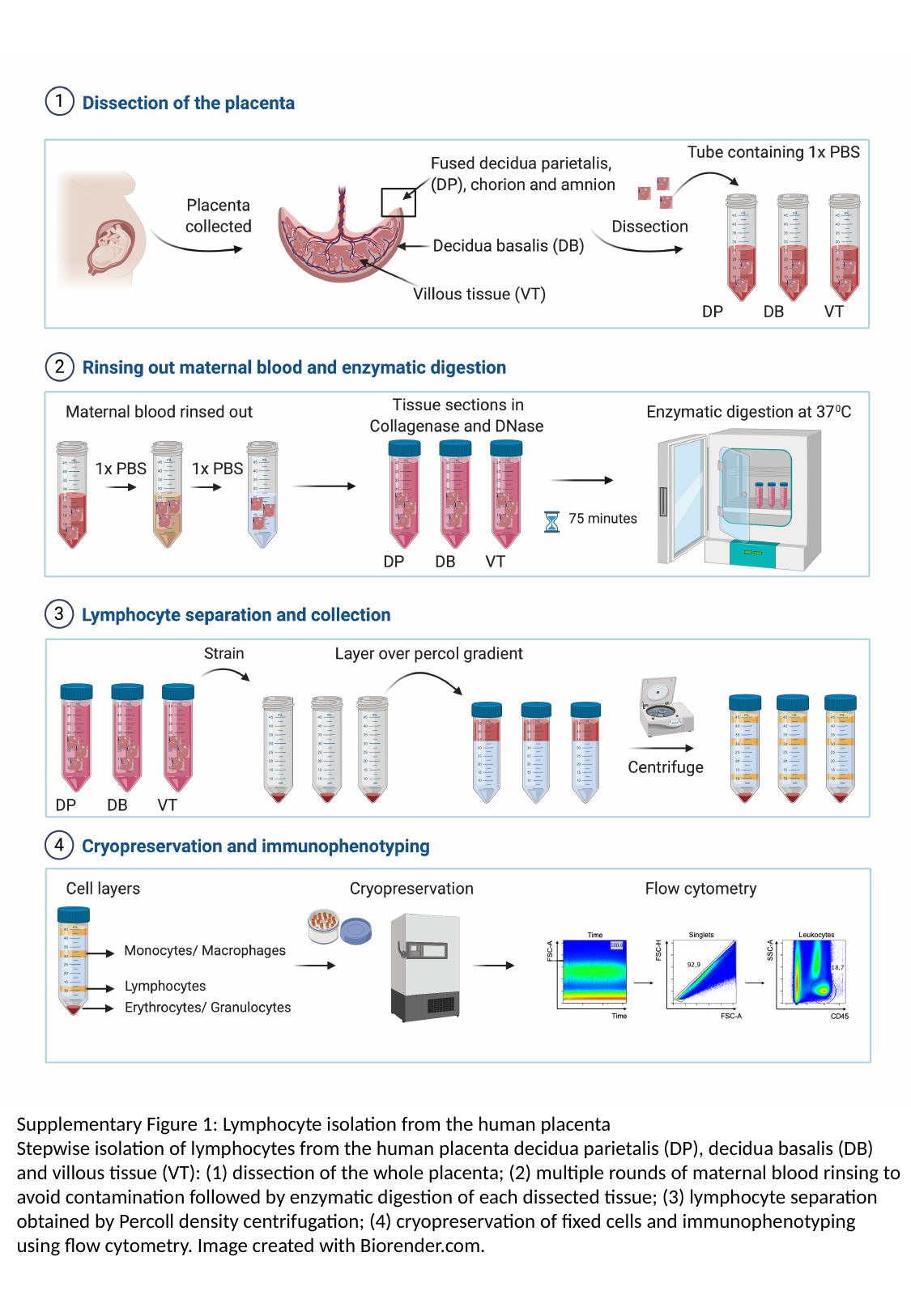

Supplementary Figure 1: Lymphocyte isolation from the human placenta
Stepwise isolation of lymphocytes from the human placenta decidua parietalis (DP), decidua basalis (DB) and villous tissue (VT): (1) dissection of the whole placenta; (2) multiple rounds of maternal blood rinsing to avoid contamination followed by enzymatic digestion of each dissected tissue; (3) lymphocyte separation obtained by Percoll density centrifugation; (4) cryopreservation of fixed cells and immunophenotyping using flow cytometry. Image created with Biorender.com.

### Slide 5
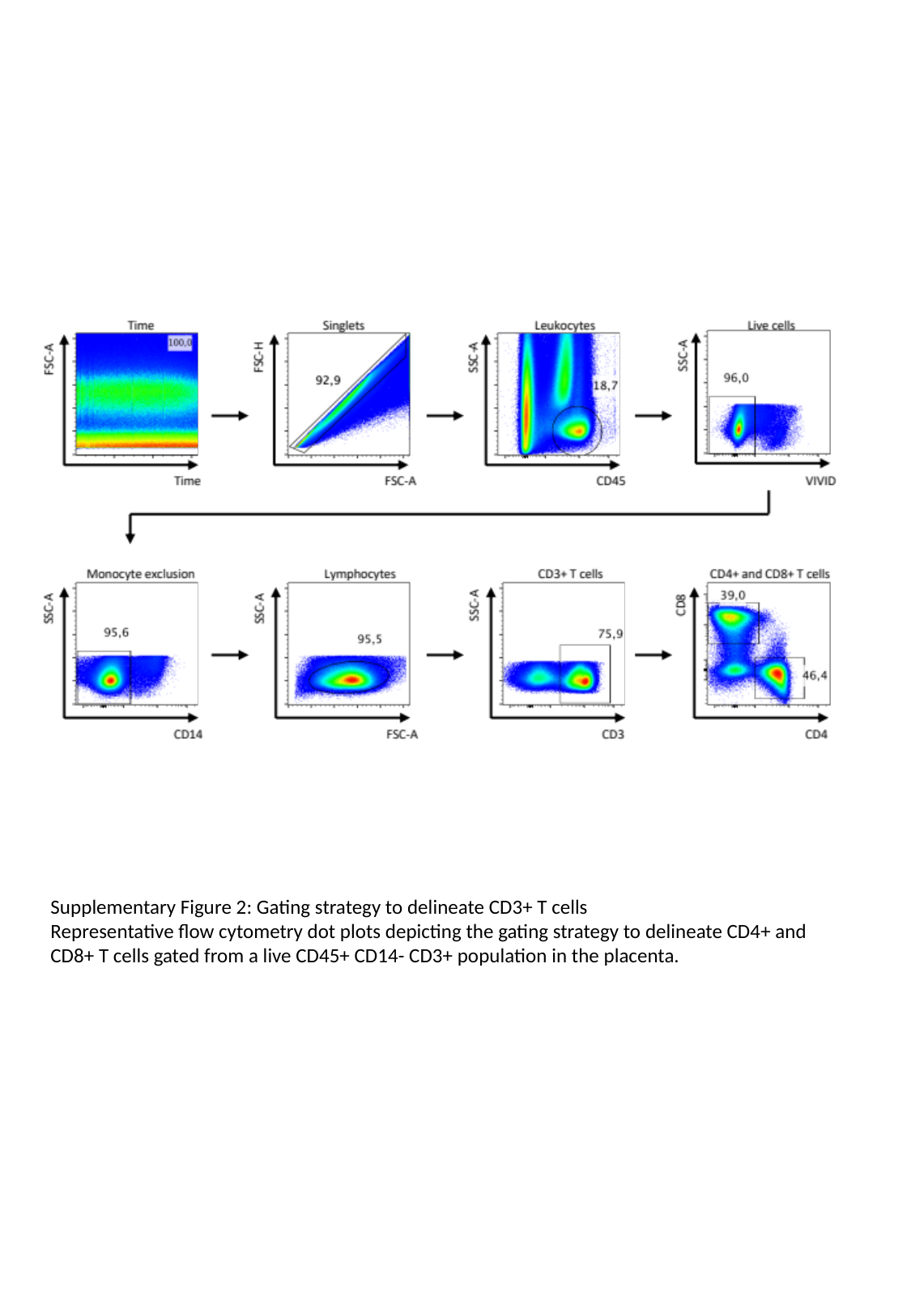

Supplementary Figure 2: Gating strategy to delineate CD3+ T cells
Representative flow cytometry dot plots depicting the gating strategy to delineate CD4+ and CD8+ T cells gated from a live CD45+ CD14- CD3+ population in the placenta.

### Slide 6
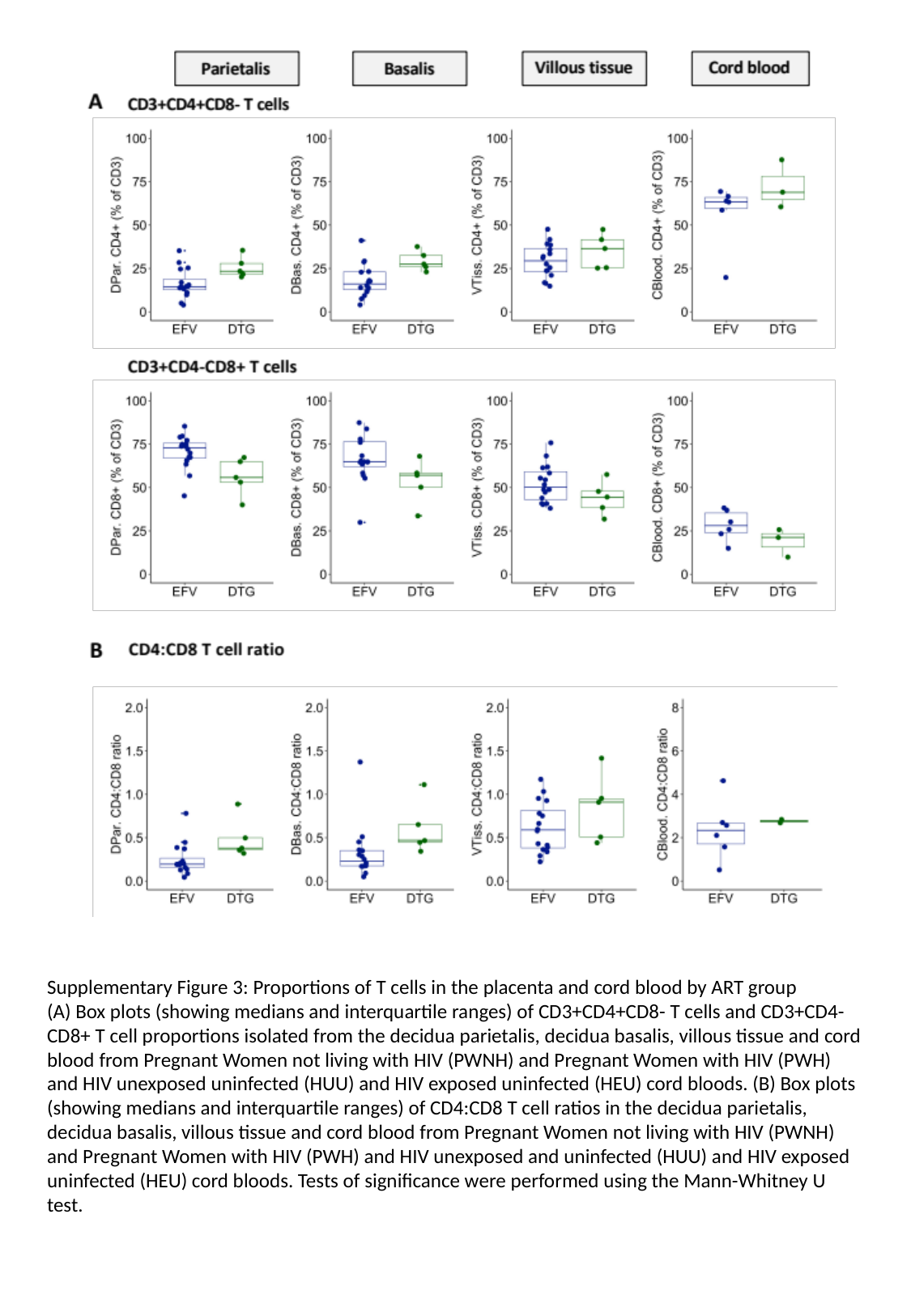

Supplementary Figure 3: Proportions of T cells in the placenta and cord blood by ART group
(A) Box plots (showing medians and interquartile ranges) of CD3+CD4+CD8- T cells and CD3+CD4-CD8+ T cell proportions isolated from the decidua parietalis, decidua basalis, villous tissue and cord blood from Pregnant Women not living with HIV (PWNH) and Pregnant Women with HIV (PWH) and HIV unexposed uninfected (HUU) and HIV exposed uninfected (HEU) cord bloods. (B) Box plots (showing medians and interquartile ranges) of CD4:CD8 T cell ratios in the decidua parietalis, decidua basalis, villous tissue and cord blood from Pregnant Women not living with HIV (PWNH) and Pregnant Women with HIV (PWH) and HIV unexposed and uninfected (HUU) and HIV exposed uninfected (HEU) cord bloods. Tests of significance were performed using the Mann-Whitney U test.

### Slide 7
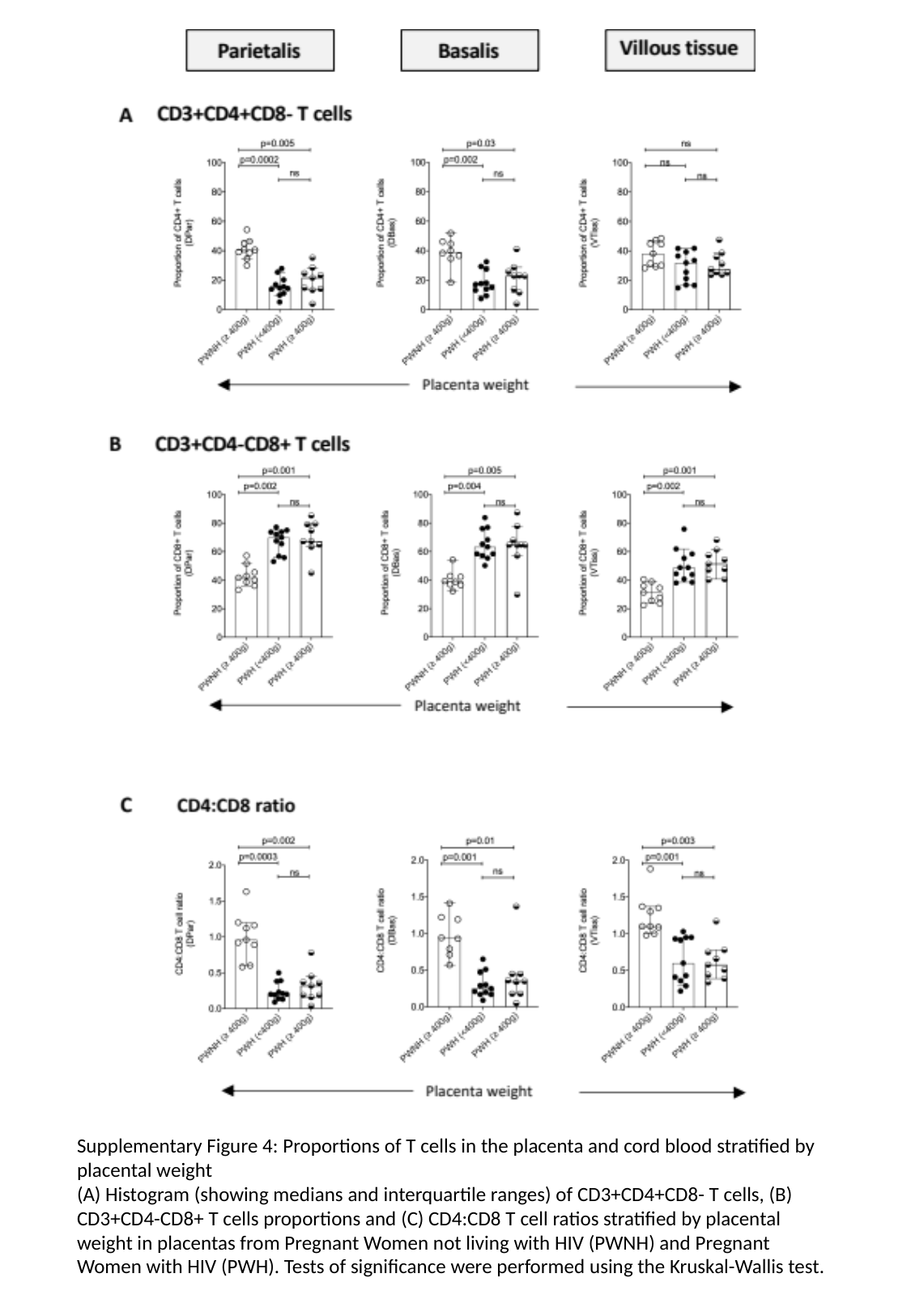

Supplementary Figure 4: Proportions of T cells in the placenta and cord blood stratified by placental weight
(A) Histogram (showing medians and interquartile ranges) of CD3+CD4+CD8- T cells, (B) CD3+CD4-CD8+ T cells proportions and (C) CD4:CD8 T cell ratios stratified by placental weight in placentas from Pregnant Women not living with HIV (PWNH) and Pregnant Women with HIV (PWH). Tests of significance were performed using the Kruskal-Wallis test.

### Slide 8
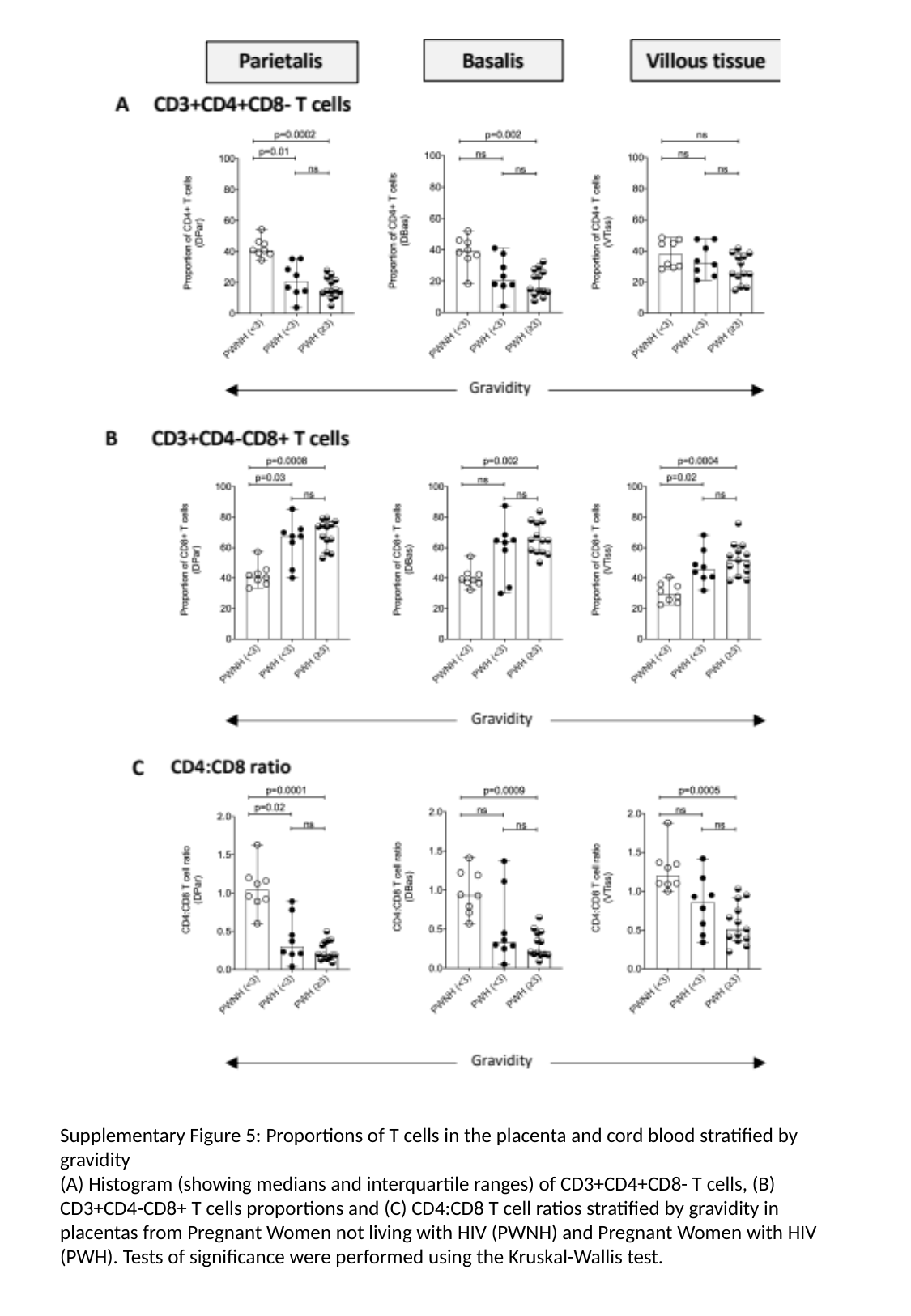

Supplementary Figure 5: Proportions of T cells in the placenta and cord blood stratified by gravidity
(A) Histogram (showing medians and interquartile ranges) of CD3+CD4+CD8- T cells, (B) CD3+CD4-CD8+ T cells proportions and (C) CD4:CD8 T cell ratios stratified by gravidity in placentas from Pregnant Women not living with HIV (PWNH) and Pregnant Women with HIV (PWH). Tests of significance were performed using the Kruskal-Wallis test.

### Slide 9
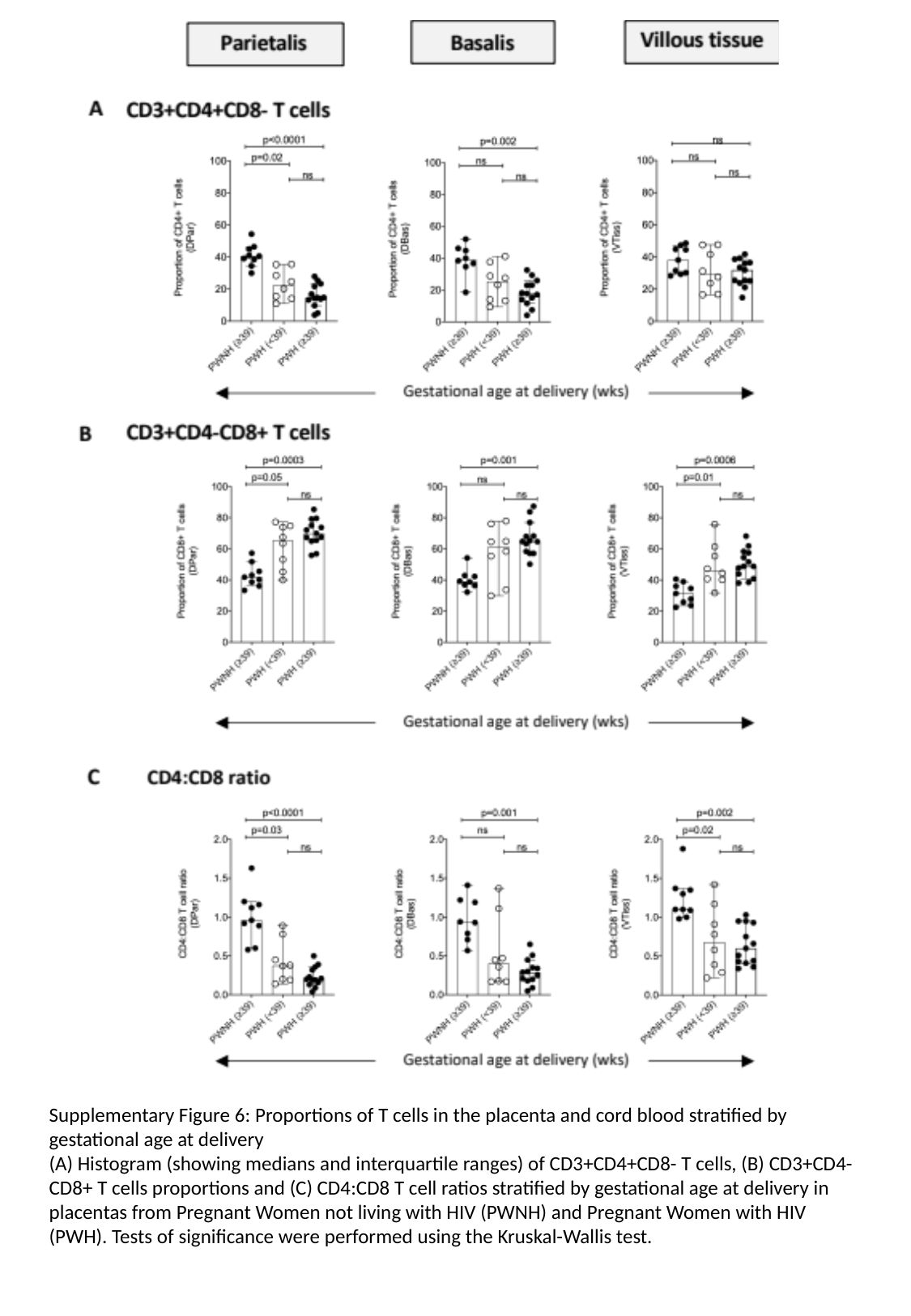

Supplementary Figure 6: Proportions of T cells in the placenta and cord blood stratified by gestational age at delivery
(A) Histogram (showing medians and interquartile ranges) of CD3+CD4+CD8- T cells, (B) CD3+CD4-CD8+ T cells proportions and (C) CD4:CD8 T cell ratios stratified by gestational age at delivery in placentas from Pregnant Women not living with HIV (PWNH) and Pregnant Women with HIV (PWH). Tests of significance were performed using the Kruskal-Wallis test.

### Slide 10
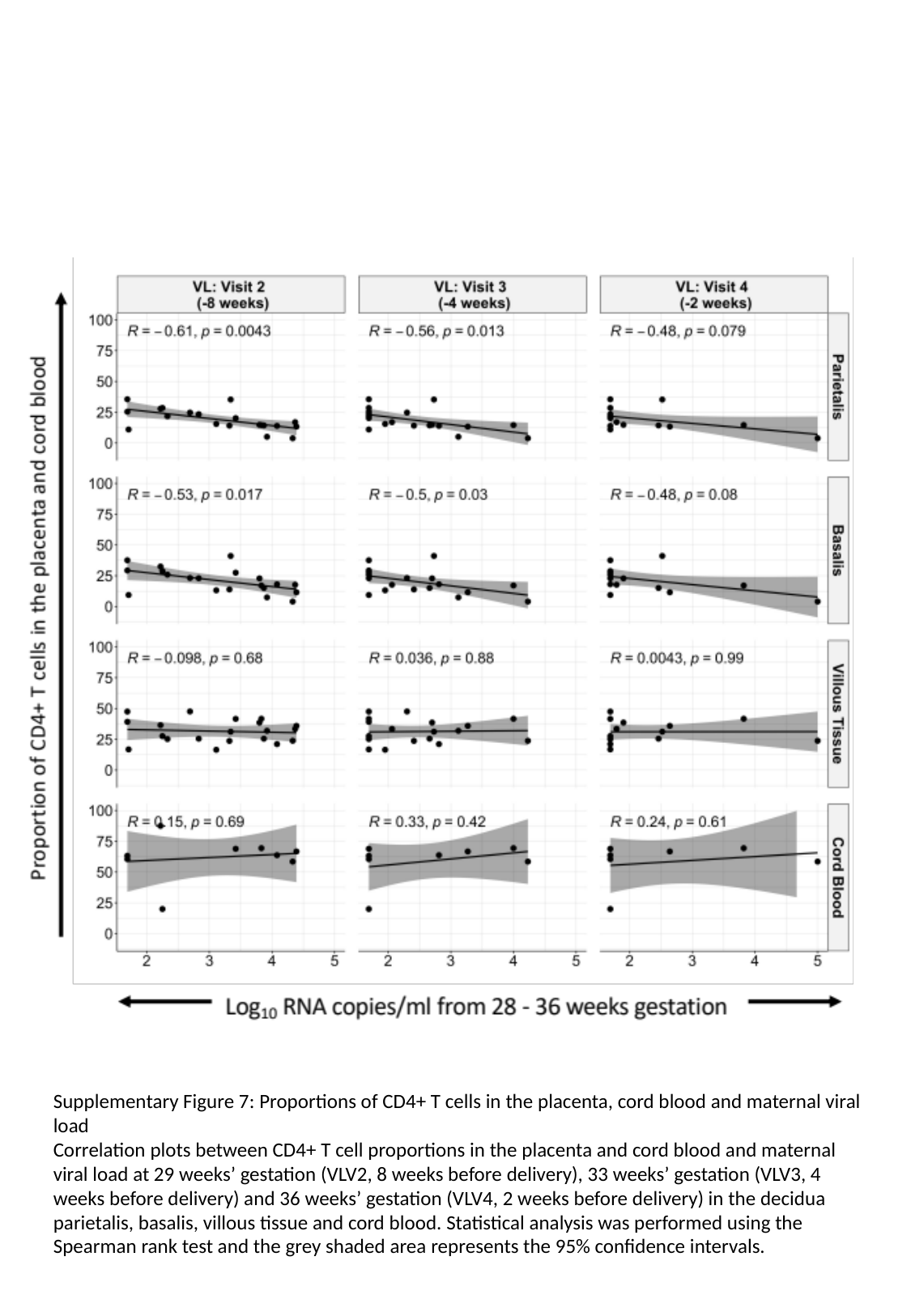

Supplementary Figure 7: Proportions of CD4+ T cells in the placenta, cord blood and maternal viral load
Correlation plots between CD4+ T cell proportions in the placenta and cord blood and maternal viral load at 29 weeks’ gestation (VLV2, 8 weeks before delivery), 33 weeks’ gestation (VLV3, 4 weeks before delivery) and 36 weeks’ gestation (VLV4, 2 weeks before delivery) in the decidua parietalis, basalis, villous tissue and cord blood. Statistical analysis was performed using the Spearman rank test and the grey shaded area represents the 95% confidence intervals.

### Slide 11
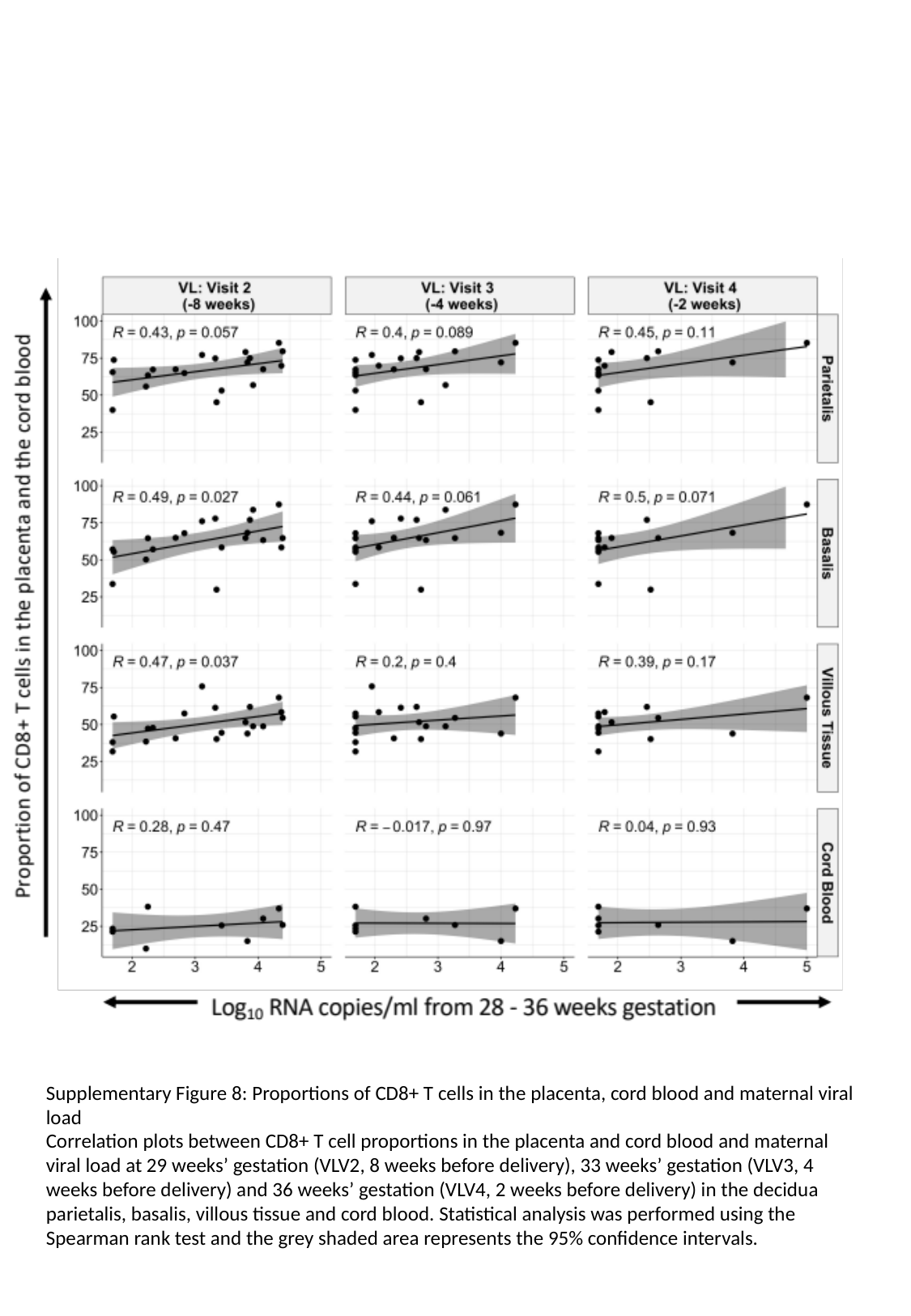

Supplementary Figure 8: Proportions of CD8+ T cells in the placenta, cord blood and maternal viral load
Correlation plots between CD8+ T cell proportions in the placenta and cord blood and maternal viral load at 29 weeks’ gestation (VLV2, 8 weeks before delivery), 33 weeks’ gestation (VLV3, 4 weeks before delivery) and 36 weeks’ gestation (VLV4, 2 weeks before delivery) in the decidua parietalis, basalis, villous tissue and cord blood. Statistical analysis was performed using the Spearman rank test and the grey shaded area represents the 95% confidence intervals.

### Slide 12
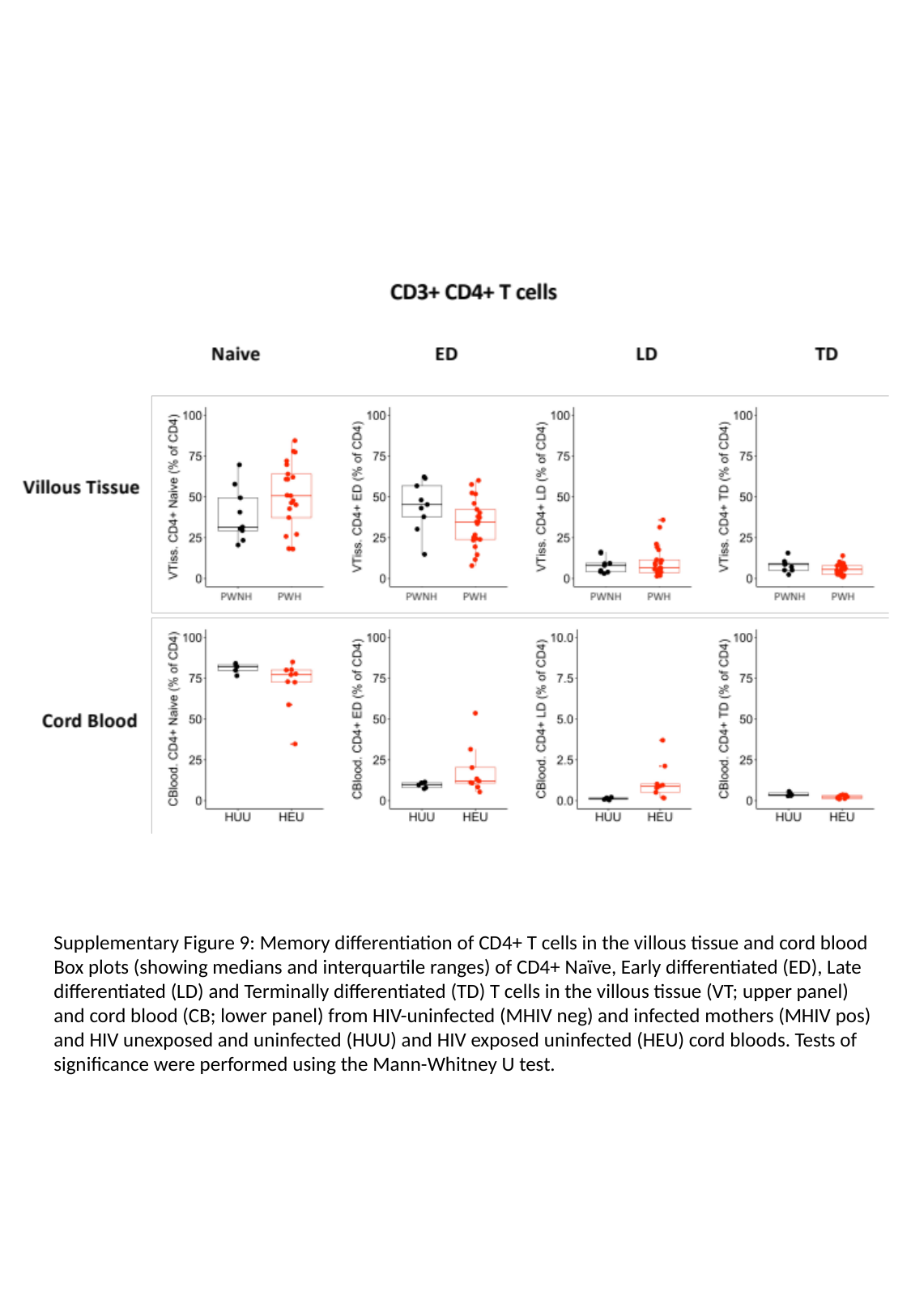

Supplementary Figure 9: Memory differentiation of CD4+ T cells in the villous tissue and cord blood
Box plots (showing medians and interquartile ranges) of CD4+ Naïve, Early differentiated (ED), Late differentiated (LD) and Terminally differentiated (TD) T cells in the villous tissue (VT; upper panel) and cord blood (CB; lower panel) from HIV-uninfected (MHIV neg) and infected mothers (MHIV pos) and HIV unexposed and uninfected (HUU) and HIV exposed uninfected (HEU) cord bloods. Tests of significance were performed using the Mann-Whitney U test.

### Slide 13
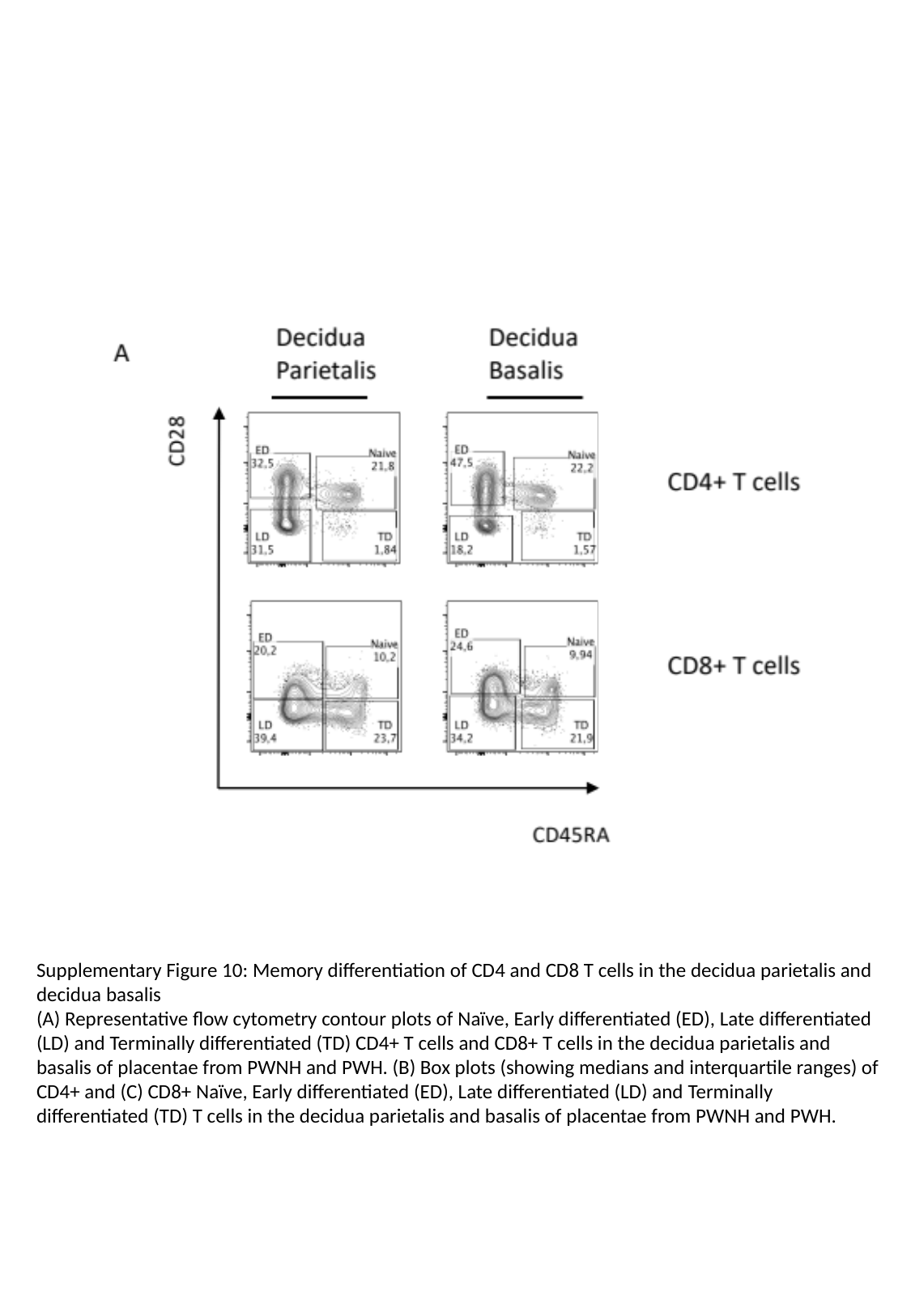

Supplementary Figure 10: Memory differentiation of CD4 and CD8 T cells in the decidua parietalis and decidua basalis
(A) Representative flow cytometry contour plots of Naïve, Early differentiated (ED), Late differentiated (LD) and Terminally differentiated (TD) CD4+ T cells and CD8+ T cells in the decidua parietalis and basalis of placentae from PWNH and PWH. (B) Box plots (showing medians and interquartile ranges) of CD4+ and (C) CD8+ Naïve, Early differentiated (ED), Late differentiated (LD) and Terminally differentiated (TD) T cells in the decidua parietalis and basalis of placentae from PWNH and PWH.

### Slide 14
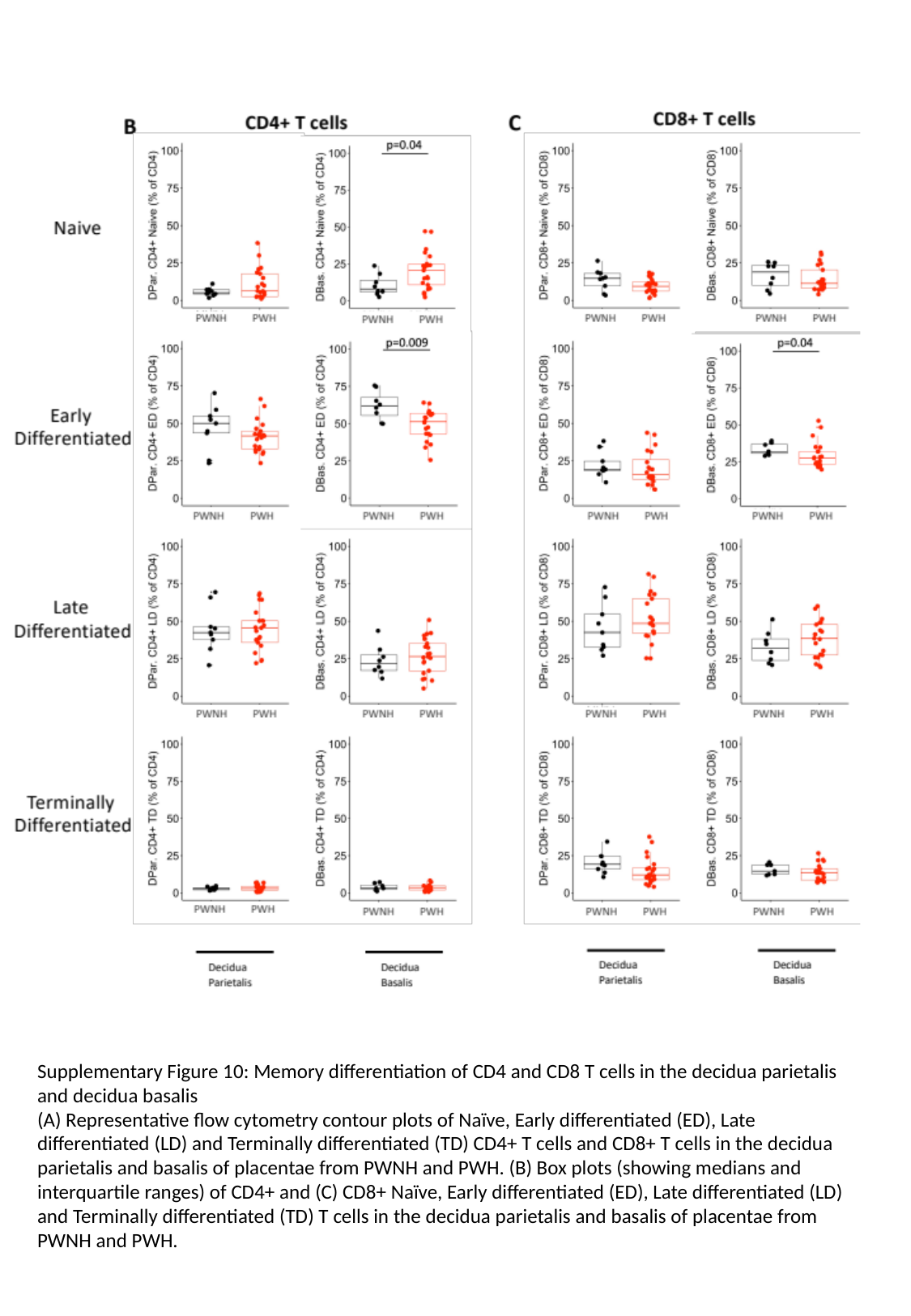

Supplementary Figure 10: Memory differentiation of CD4 and CD8 T cells in the decidua parietalis and decidua basalis
(A) Representative flow cytometry contour plots of Naïve, Early differentiated (ED), Late differentiated (LD) and Terminally differentiated (TD) CD4+ T cells and CD8+ T cells in the decidua parietalis and basalis of placentae from PWNH and PWH. (B) Box plots (showing medians and interquartile ranges) of CD4+ and (C) CD8+ Naïve, Early differentiated (ED), Late differentiated (LD) and Terminally differentiated (TD) T cells in the decidua parietalis and basalis of placentae from PWNH and PWH.

### Slide 15
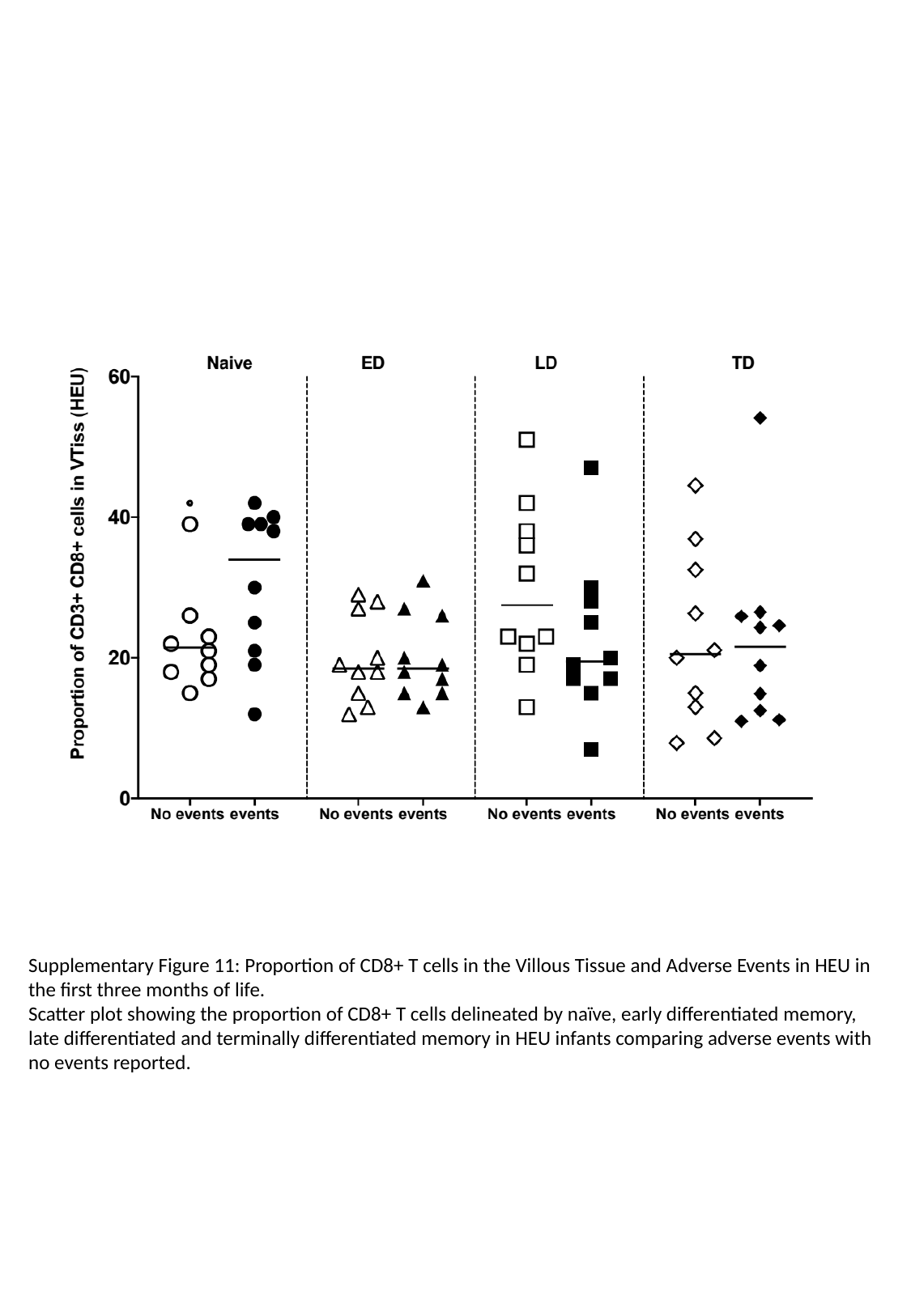

Supplementary Figure 11: Proportion of CD8+ T cells in the Villous Tissue and Adverse Events in HEU in the first three months of life.
Scatter plot showing the proportion of CD8+ T cells delineated by naïve, early differentiated memory, late differentiated and terminally differentiated memory in HEU infants comparing adverse events with no events reported.
